# Optical compression of multimodal clinical data into a unified visual memory for generative trajectory forecasting

**DOI:** 10.64898/2026.09.03.26361736

**Authors:** Zongheng Guo, Bo Jin, Zhi Han, Takumi Ichikawa, Naoto Ozawa, Xuefeng B. Ling

## Abstract

**Importance:** Current clinical artificial intelligence is bottlenecked by highly specialized, fragmented models that reduce complex, multimodal data into isolated scalar predictions. By reframing multimodal perception as a pure vision problem, optical compression into a universal visual operating system can resolve internal data interaction bottlenecks and shift the paradigm toward continuous, generative clinical forecasting.

**Objective:** To develop and internally validate Clinical Visual Memory, a generalist visual foundation model utilizing high-fidelity optical compression to fuse five heterogeneous intensive care unit (ICU) data modalities into a unified visual-token space, acting as a generative navigation system for critical-care trajectories.

**Design, Setting, and Participants:** Retrospective modeling study using MIMIC-IV adult ICU encounters. After excluding stays shorter than 24 hours, 54,551 patients, 68,546 hospital admissions, and 74,829 ICU stays comprised the eligible cohort. Strict patient-level partitioning prevented cross-split leakage.

**Methods:** Structured electronic health record (EHR) data, vital signs, 10-second electrocardiogram (ECG) waveforms, chest radiographs, and clinical notes were rendered as 2D images and encoded by a single frozen DINOv2 vision transformer. Through modality-aware latent-query cross-attention, these highly heterogeneous sources were optically compressed into a shared 1024-dimensional Clinical Visual Memory. To establish a universal output interface, this memory conditioned an conditional instruction-tuned image generator to decode eight-domain deterioration trajectories across 3- to 48-hour horizons. Visual compression fidelity was evaluated by reducing retained source-pixel area to 1%, and critical transitions were mapped using event-specific projected-axis geometry.

**Results:** Operating as a generalist foundation, Clinical Visual Memory achieved AUROCs of 0.852 (95% CI, 0.821-0.882) for 48-hour mortality, 0.699 (0.673-0.725) for incident acute kidney injury (AKI), 0.723 (0.688-0.757) for high Sequential Organ Failure Assessment (SOFA), and 0.742 (0.726-0.757) for alive ICU discharge. Crucially, resolving the token bottleneck via an 8-fold reduction in source-pixel area preserved 98.9% of the uncompressed 48-hour mortality AUROC. Generative image-out decoding maintained clinical meaning, and trajectory geometry functioned as a proactive navigation system, yielding median warning lead times of 34.8 hours (IQR, 17.5-43.3) before death, 14.6 hours (6.5-33.2) before AKI, 15.0 hours (6.0-27.0) before high SOFA, and 20.8 hours (10.2-36.0) before recovery, with low false-alert burdens (0.040-0.109 per patient-day).

**Conclusions and Relevance:** Clinical Visual Memory demonstrates that optical compression can successfully unify highly heterogeneous clinical data into a single, high-fidelity visual-token representation. By functioning as a continuous clinical navigation system rather than a discrete alert generator, this framework lays the architectural foundation for a generalist, generative visual operating system in critical care medicine. External and prospective validation are required before clinical deployment.

**KEY POINTS:** *Question:* Can optical compression of heterogeneous, multimodal intensive care data into a unified visual-token memory enable a generalist generative framework for continuous clinical forecasting?

*Findings:* In MIMIC-IV, five clinical modalities were rendered as images and fused via a shared frozen DINOv2 transformer into a 1024-dimensional Clinical Visual Memory. This unified architecture supported robust multitask prediction, achieving AUROCs of 0.852 for 48-hour mortality, 0.699 for incident AKI, 0.723 for high SOFA, and 0.742 for alive ICU discharge. Crucially, an 8-fold reduction in source-pixel area preserved 98.9% of the uncompressed mortality discrimination, validating high-fidelity optical compression. Generative image-out decoding successfully translated this memory into actionable clinical trajectories, yielding median warning lead times of 34.8 hours before death, 14.6 hours before AKI, 15.0 hours before high SOFA, and 20.8 hours before alive ICU discharge.

*Meaning:* By resolving multimodal data bottlenecks through high-fidelity visual compression, this framework shifts clinical AI from fragmented, task-specific alerts to a unified, generalist visual operating system. It demonstrates that complex patient states can be continuously navigated and generatively forecasted via a single interpretable visual-token memory.

## INTRODUCTION

Critical care requires continuous state estimation to navigate a patient’s trajectory toward crisis or recovery. Yet, clinical artificial intelligence remains bottlenecked by highly specialized, fragmented models that reduce complex, multimodal data into isolated scalar predictions [1–3]. Established scoring systems like APACHE II, SAPS II, and SOFA similarly compress acute illness into static indices rather than persistent, queryable representations [4–6].

This fragmentation prevents AI from acting as a continuous clinical navigation system, exposing the absence of a unified patient-state substrate. This fragmentation originates at the input layer. Structured electronic health records, irregular time series, waveforms, radiographs, and clinical narratives differ fundamentally in dimensionality, semantics, and missingness. While multimodal biomedical frameworks demonstrate that combining these sources improves prediction [7–9], they typically rely on modality-specific feature extractors fused at a late stage. This siloed approach exacerbates internal token bottlenecks and produces narrow, endpoint-specific alerts prone to calibration drift and alert fatigue [10]. A truly generalist architecture must decouple the core representation of the patient from the specific clinical question being asked.

The transition toward foundation models suggests a paradigm shift: learning a highly compressed, reusable representation adaptable to multiple downstream tasks [11, 12–14]. However, unifying longitudinal, whole-patient information into a single compressible and machine-decodable substrate remains a formidable challenge. We hypothesize that visual rendering provides the optimal universal interface for this substrate. Medicine inherently expresses meaning through spatial arrangements— temporal morphology in waveforms, anatomy in radiographs, multivariate state in heatmaps, and structured hierarchy in documents. Vision transformers allow these disparate canvases to be represented as standard token sequences [15], while cross-attention architectures distill massive, heterogeneous inputs into a compact latent array [16].

Crucially, a pure visual substrate resolves the mechanistic challenge of information density. Long clinical histories rapidly exceed the context windows of longitudinal electronic health record models, forcing aggressive truncation [17]. Inspired by recent breakthroughs demonstrating that optical 2D mapping can radically compress long contexts [18], we introduce optical compression as a mechanism to resolve the multimodal token crisis in medicine. Furthermore, framing perception as image generation establishes a unified, generalist interface for complex tasks [19]. By converting heterogeneous clinical sources into a compatible visual-token grammar, disparate modalities can be aggressively compressed while retaining their clinical identity.

Here, we present Clinical Visual Memory, an end-to-end framework that optically compresses five disparate intensive care modalities into a unified visual-token memory (Figure 1). Rather than generating discrete alerts, this generalist visual operating system functions as a continuous clinical navigation system, decoding highly compressed latent representations into generative, multiorgan trajectories to accurately forecast deterioration and recovery.

**Figure 1.**
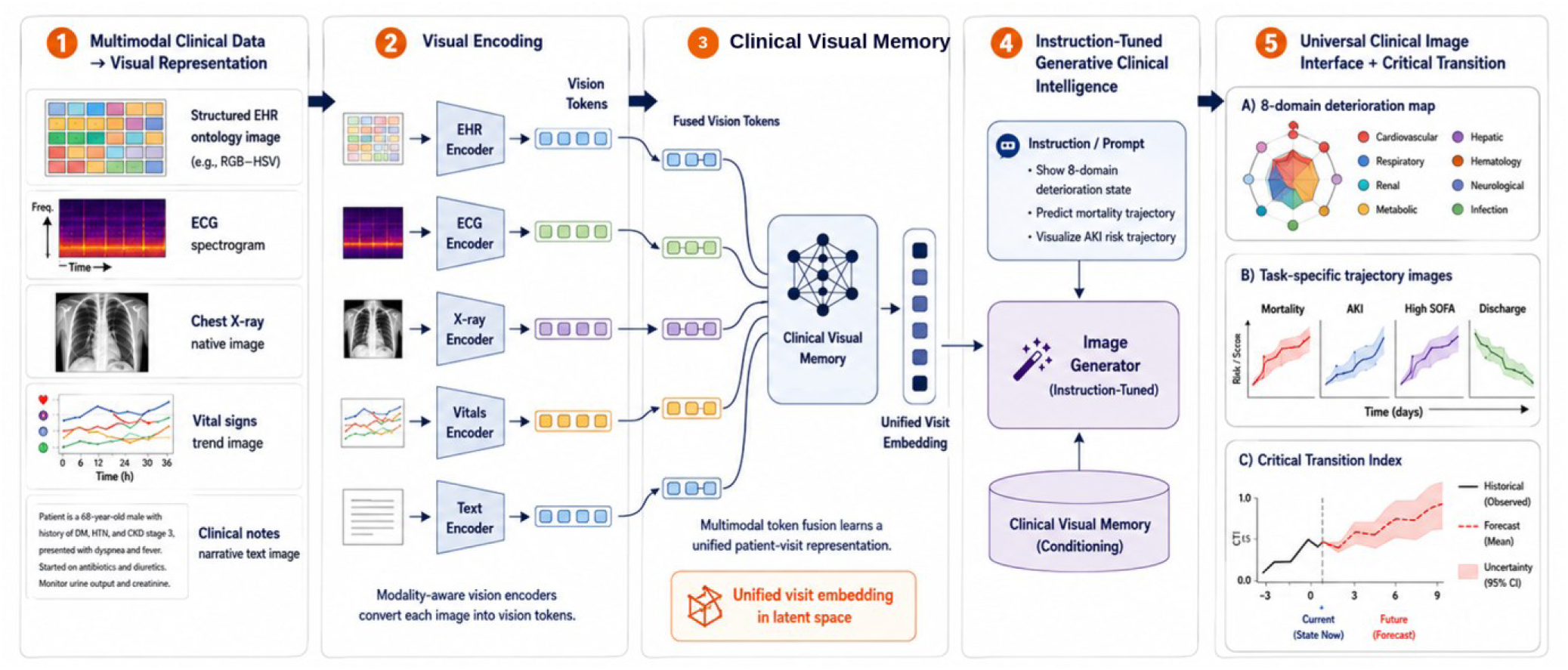
Clinical Visual Memory architecture and analytical scope. Five ICU modalities are converted to standardized visual representations and processed using the same frozen DINOv2 ViT-L/14 weights. Modality-specific projections, source embeddings, explicit availability masks, and latent-query fusion produce a 1024-dimensional Clinical Visual Memory that supports direct 48-hour prediction and task-conditioned image-out. The source-image compression experiment intervenes before encoding while holding the restored canvas, encoder geometry, and patch-token count fixed. Critical-transition lead times are derived from a separate eight-domain observed-state geometry and are not outputs of Clinical Visual Memory or image-out. Abbreviations: AKI, acute kidney injury; CVM, Clinical Visual Memory; EHR, electronic health record; ECG, electrocardiogram; ICU, intensive care unit; SOFA, Sequential Organ Failure Assessment; STFT, short-time Fourier transform.

## METHODS

### Study design, data sources, and cohort

To develop and internally validate this generalist generative forecasting framework, we conducted a retrospective, fixed-landmark modeling study using linked, deidentified MIMIC-IV critical-care data, MIMIC-IV-ECG waveforms, MIMIC-CXR radiographs, and MIMIC-IV-Note clinical narratives from Beth Israel Deaconess Medical Center [20–23]. The ICU stay was the primary modeling unit, and the prespecified prediction landmark was 24 hours after ICU admission. Model inputs were restricted to information accrued during hours 0–24, and forecast outcomes were defined strictly after the landmark according to the outcome-specific risk-set criteria described below.

The source extraction contained 65,366 patients, 85,242 hospital admissions, and 94,458 ICU stays; no patient was younger than 18 years. Of these, 19,629 ICU stays (20.8%) ended before the 24-hour landmark and therefore did not provide an eligible hour-24 prediction occasion. Their exclusion defined the primary landmark population and was not a complete-case restriction based on missing measurements or modalities. The resulting cohort comprised 54,551 patients, 68,546 hospital admissions, and 74,829 ICU stays in which the patient remained under ICU care at hour 24. Among stays reaching the landmark, unavailable measurements or modalities were represented using explicit availability masks [if accurate]. Subsequent temporal and modality-linkage quality control excluded no additional patients or stays.

The chronologically first eligible ICU stay for each patient was used for descriptive cohort characteristics; modeling analyses used all eligible stays meeting the applicable outcome-specific criteria (Figure 2A). Patients were randomly assigned using a fixed seed to training, validation, and held-out test partitions in an 80:10:10 ratio. All eligible ICU stays belonging to an individual patient were assigned to the same partition to prevent cross-split leakage. No formal sample-size calculation was performed; all eligible data were used. Reporting followed STROBE and TRIPOD+AI [24,25].

**Figure 2.**
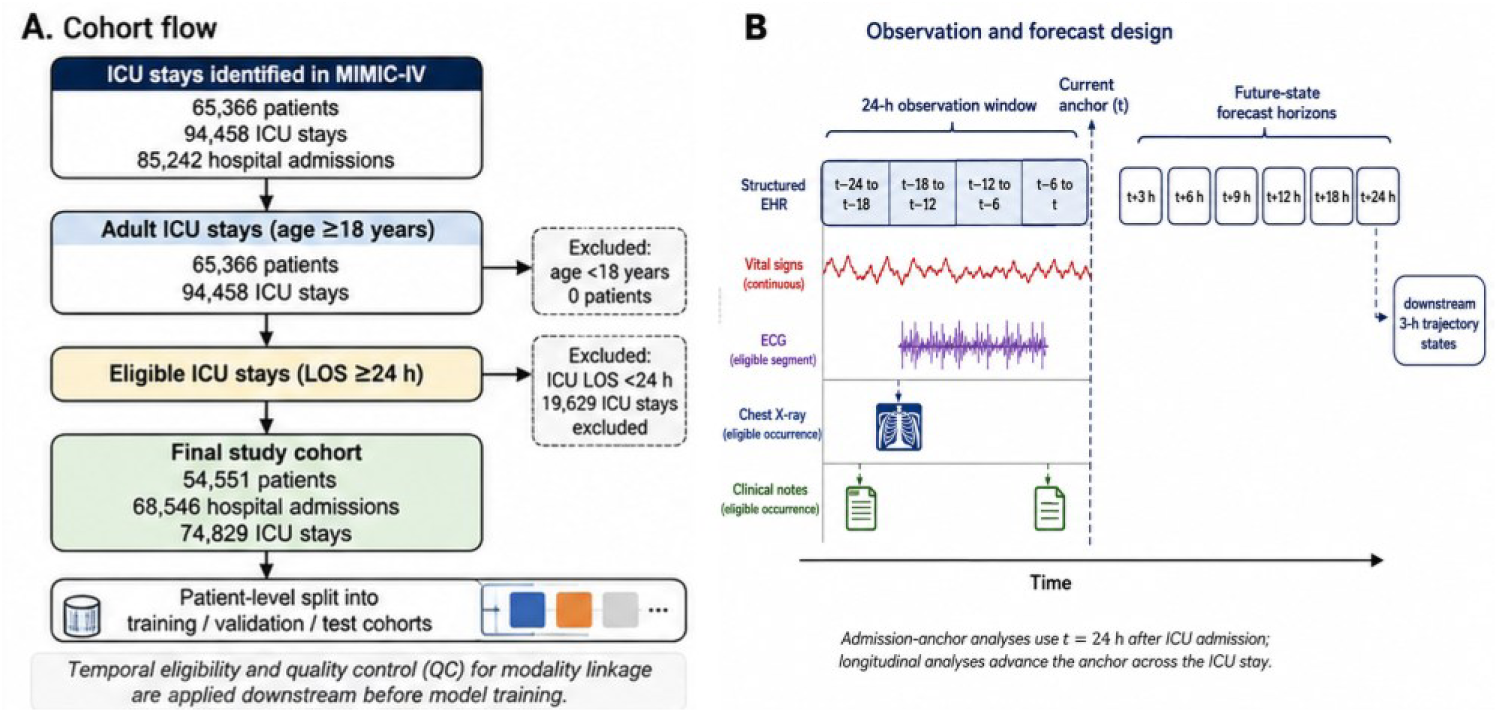
Cohort construction and temporal design of the Clinical Visual Memory study. (A) The MIMIC-IV source extraction contained 65,366 patients, 85,242 hospital admissions, and 94,458 ICU stays. Exclusion of 19,629 stays shorter than 24 hours left 54,551 patients, 68,546 hospital admissions, and 74,829 ICU stays before modality-linkage and temporal quality control, which excluded no additional patients or stays, confirming the final analytic cohort at N=54,551 patients (54,551 index ICU stays), as shown in the updated Panel A. Patient-level partitioning prevents the same patient from crossing training, validation, and test sets. (B) At each prediction anchor t, the preceding 24 hours form the observation window. Structured EHR data are summarized in four 6-hour bins; vital signs retain continuous timing; and eligible ECG, chest-radiograph, and note occurrences are aligned within the same window. Primary future-state generation uses sequential 3-hour horizons, whereas the held-out classification tasks use 48-hour outcome windows.

### Prediction anchor and clinical outcomes

For the admission-anchored generative forecasting analyses, the anchor was 24 hours after ICU admission. Data from the preceding 24 hours constituted the observation window (Figure 2B). Structured EHR measurements were summarized in four consecutive 6-hour bins. Vital signs retained their recorded sampling times, and temporally eligible electrocardiograms, chest radiographs, and clinical notes were aligned to the same observation window. Four binary outcomes were evaluated during the 48 hours after the anchor: in-hospital mortality, incident acute kidney injury (AKI), first high Sequential Organ Failure Assessment (SOFA) state, and alive ICU discharge. In-hospital mortality was defined as death within 48 hours after the anchor and before hospital discharge. Incident AKI was the first post-anchor event meeting Kidney Disease: Improving Global Outcomes stage 1 or higher according to serum-creatinine or urine-output criteria [26]. High SOFA was the first rolling 24-hour SOFA score of 6 or greater after the anchor [6]. SOFA was recomputed at 3-hour intervals using its six standard organ-system components and was considered evaluable when at least four components were available. High SOFA was treated as an organ-dysfunction endpoint and not as synonymous with sepsis [27]. Alive ICU discharge was discharge from the ICU within the prediction window among patients alive at ICU discharge, operationalized as an ICU outtime preceding any recorded death time or occurring in the absence of a recorded death time. At-risk populations were constructed separately for each outcome. Stays were excluded from a task if the corresponding event had occurred at or before the anchor, including prevalent AKI, SOFA of 6 or greater, or death. Stays without observable post-anchor follow-up were also excluded.

### Multimodal visual input construction for optical compression

To prepare highly heterogeneous clinical data for optical compression, five clinical modalities were converted into standardized visual canvases: structured EHR data, vital-sign trajectories, electrocardiography, chest radiography, and clinical notes. Structured-data extraction and temporal alignment followed established MIMIC preprocessing principles [28]. Structured measurements were represented as a visit-level RGB/HSV ontology canvas containing 38 clinical concepts organized into 10 semantic systems. Static variables were repeated across the four 6-hour bins, while nonstatic variables were aggregated using prespecified rules according to clinical directionality. Values were mapped to bounded abnormality scores from 0 to 1. Hue denoted semantic system, increasing saturation denoted increasing abnormality, and decreasing brightness denoted increasing severity. Missing measurements were assigned a reserved neutral-gray state and recorded in a separate missingness mask. The resulting matrix was expanded to a 224-by-224-pixel RGB canvas. Detailed variable mappings and ranges are provided in Supplementary Table 1. Vital-sign images comprised seven stacked trajectories (heart rate, respiratory rate, peripheral oxygen saturation, systolic blood pressure, diastolic blood pressure, mean arterial pressure, and temperature) plotted at their recorded times over the observation window on an 896-by-896-pixel canvas. Missing intervals remained unobserved rather than being bridged.

Electrocardiographic waveforms were obtained from MIMIC-IV-ECG [21]. Recordings comprised 10-second, 12-lead diagnostic electrocardiograms sampled at 500 Hz. Lead I was robustly standardized and transformed into a short-time Fourier spectrogram using 256-sample windows with 192-sample overlap. Frequencies above 45 Hz were excluded. Log power was clipped to the within-image 2nd and 98th percentiles, normalized to the range from 0 to 1, and rendered as an 896-by-896-pixel image. Chest radiographs from MIMIC-CXR [22] underwent orientation correction when required and robust intensity normalization to the 0.5th and 99.5th percentiles. Images were resized to 896 by 896 pixels and represented as grayscale information replicated across three channels. Clinical notes from MIMIC-IV-Note [23] were ordered chronologically, concatenated, deterministically truncated at 12,000 characters, and rendered using fixed typography onto an 896-by-896-pixel document image.

### Optical compression via shared visual encoding and availability-aware fusion

To resolve multimodal data bottlenecks, all five image types were processed by the same frozen DINOv2 ViT-L/14 vision backbone [15,29] (Figure 3A; Table 2). The final hidden-state sequence for each modality was retained, including the class token and full patch-token sequence; model-specific register tokens were excluded. Each retained visual token was 1024-dimensional. Encoder identity, hidden dimension, token count, numerical finiteness, and class/patch-token consistency were checked before fusion. Each modality was mapped into a common 1024-dimensional fusion space through a modality-specific LayerNorm–linear–GELU–LayerNorm projection (Figure 3B). A learned modality embedding was added to preserve source identity. A five-element binary vector indicated whether structured EHR, vital signs, electrocardiogram, chest radiograph, and clinical-note modalities were available. Tokens corresponding to unavailable modalities were hard-masked from attention rather than represented as clinical zeros, and the availability vector also conditioned the fused state. During training, observed-modality dropout was applied with probability 0.15 while ensuring that at least one observed modality remained. Tokens from available modalities were concatenated and attended to by 16 learnable latent queries (Figure 3C), following the general cross-attention principle of latent-array architectures [16]. Two fusion blocks each contained latent-to-input cross-attention, self-attention among the latent queries, and a feed-forward network with residual connections and normalization. The primary configuration used 8 attention heads and a 2048-dimensional feed-forward layer. A learned pooling query reduced the latent array to a 1024-dimensional Clinical Visual Memory:

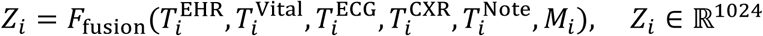

**Figure 3.**
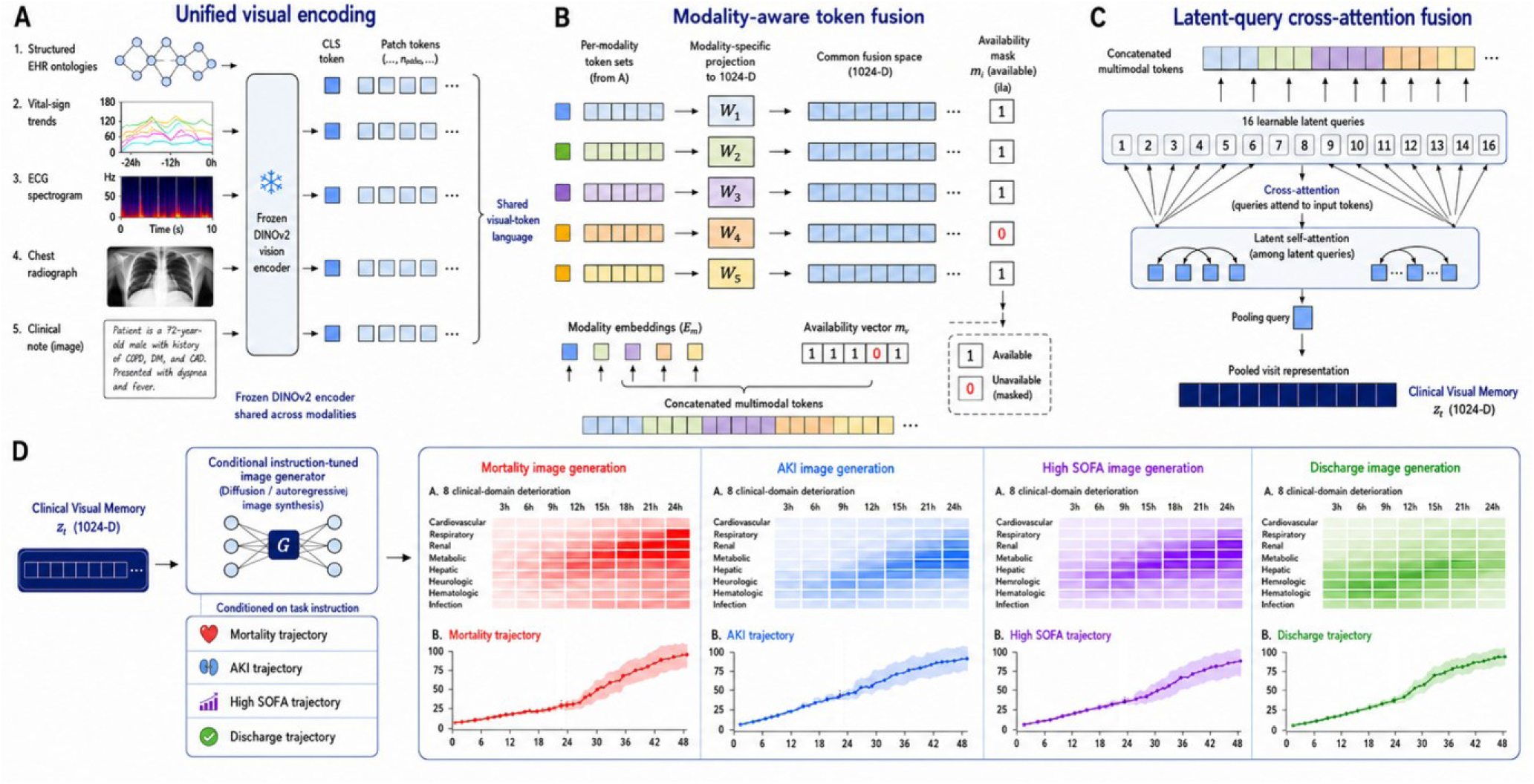
Clinical Visual Memory architecture and instruction-conditioned generative image-out. (A) Unified visual encoding: the five modality images are processed by the same frozen DINOv2 vision encoder, and class and patch tokens are retained. (B) Modality-aware token fusion: modality-specific projections map tokens into a common 1024-dimensional space, learned modality embeddings preserve source identity, and an explicit availability mask excludes unavailable modalities. (C) Latent-query cross-attention: 16 learnable latent queries cross-attend to concatenated multimodal tokens, undergo latent self-attention, and are pooled to produce the 1024-dimensional Clinical Visual Memory. (D) Instruction-conditioned generative image-out: the Clinical Visual Memory is combined with a task instruction specifying mortality, acute kidney injury, high SOFA, or alive ICU discharge trajectory and passed to a conditional instruction-tuned image generator using latent-diffusion image synthesis. Each instruction produces a task-specific clinical image containing two components: an eight-domain deterioration heatmap and the corresponding outcome trajectory.

### Multitask representation learning and comparators

The modality-specific projections, latent-query fusion blocks, pooling layer, and task-specific prediction heads were optimized jointly using the four prespecified clinical objectives. Binary outcomes were optimized using binary cross-entropy with logits and training-derived positive-class weights, utilizing AdamW optimization [30]. Two comparators were prespecified. An L2-regularized logistic-regression model used matched structured clinical information from the same observation window. A pooled-vision ablation used the same visual features as Clinical Visual Memory but replaced the latent-query optical compression fusion with a simpler pooled visual representation.

### Generative forecasting via task-conditioned image-out decoding

To complete the narrative arc from visual input to generative forecasting, the 1024-dimensional Clinical Visual Memory conditioned an image-out pathway. Ground-truth target images were constructed deterministically from observed future clinical states to ensure each output region remained machine-decodable. Each 1024-by-768-pixel generative output combined two panels: an eight-domain physiological deterioration map and an endpoint-proximity trajectory (mortality, AKI, high SOFA, or alive ICU discharge) spanning 16 sequential 3-hour horizons. Each panel type was compressed by a dedicated convolutional autoencoder into an 8-channel latent representation. Conditional U-Net diffusion components used a 1000-step cosine noise schedule, denoising diffusion implicit-model sampling, and classifier-free guidance [31–34]. The eight-domain panel was conditioned on Clinical Visual Memory, while the task-specific panel was conditioned on Clinical Visual Memory together with a learned task-identity token. The task-panel pathway utilized a two-stage, patient-anchored design: a deterministic decoder mapped the memory to a conditional-mean trajectory latent, and the diffusion model generated the normalized residual. Predetermined generative image regions were decoded algorithmically into task-specific risk or discharge-proximity estimates and compared with held-out outcomes.

### Validation of high-fidelity optical compression

To validate the robustness of the optical compression input mechanism, compression was evaluated at the source-image level (Figure 4). At each retained-area budget, images were downsampled isotropically using a modality-specific interpolation kernel and subsequently restored to their original canvas dimensions. This operation discarded spatial detail while holding encoder input dimensions and DINOv2 patch-token geometry constant. Retained source-pixel areas were 100%, 50%, 25%, 12.5%, 10%, 5%, 2%, and 1%, corresponding to nominal compression ratios of 1-, 2-, 4-, 8-, 10-, 20-, 50-, and 100-fold. Images in the 100% condition were not resampled. The compression-performance measure was 48-hour mortality AUROC for the full availability-aware fusion model. At retained-area fraction r, performance retention was calculated as

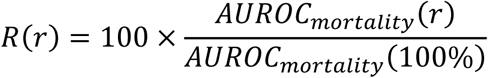

**Figure 4.**
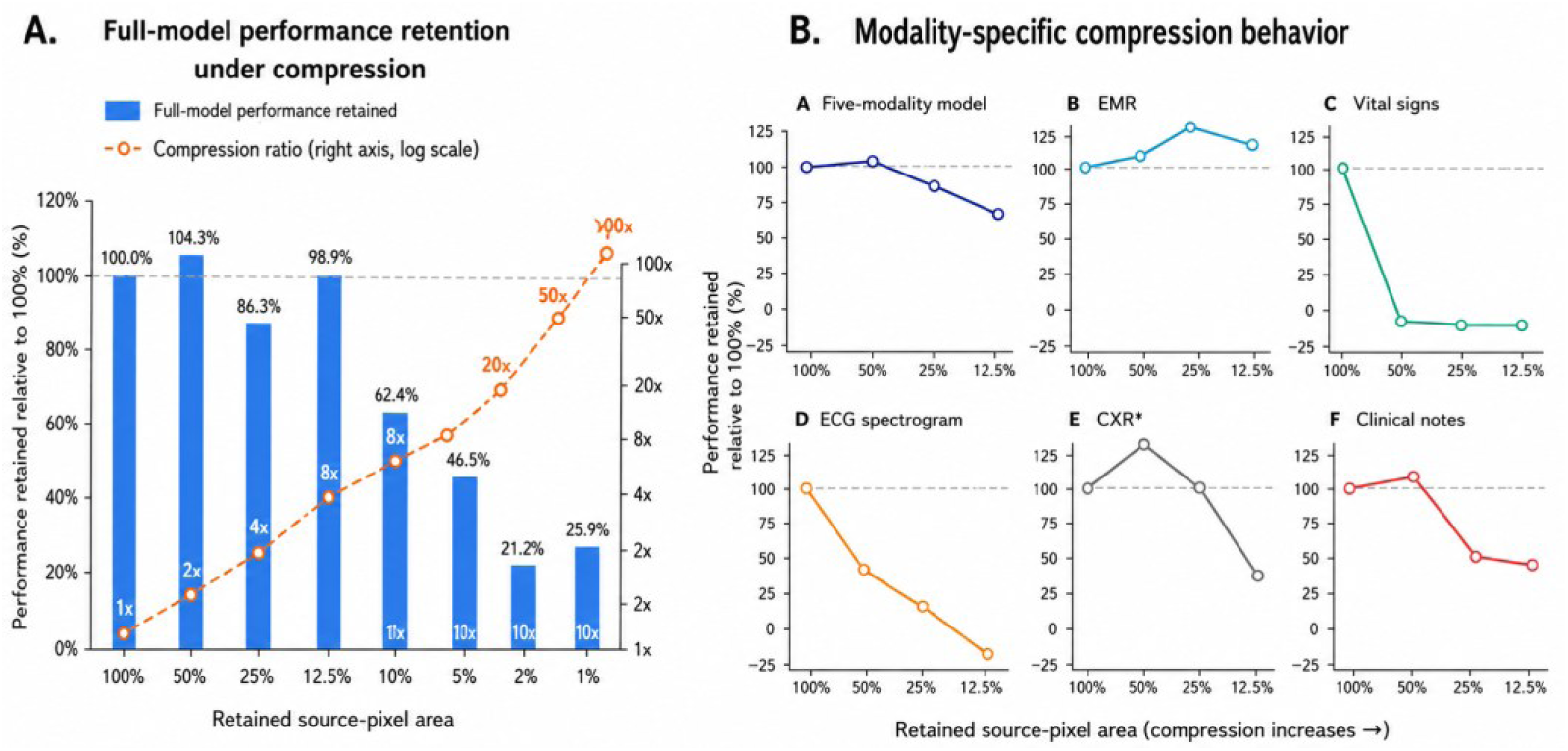
Clinical information retention under multimodal visual compression. (A) Mortality-discrimination retention for the full availability-aware fusion model is shown relative to the 100% source-pixel reference across retained source-pixel areas from 100% to 1%; the secondary axis denotes the nominal compression ratio from 1× to 100×. Retained performance is defined as 100 × *AUROC_mortality_*(*r*)/*AUROC_mortality_*(100%), where r is the retained source-pixel fraction. (B) Modality-specific compression behavior uses the same mortality-AUROC retention definition, with each modality-specific curve normalized to its own 100% source-pixel reference. The experiment measures retained source-image information rather than direct reduction of DINOv2 patch-token count.

**Figure 5.**
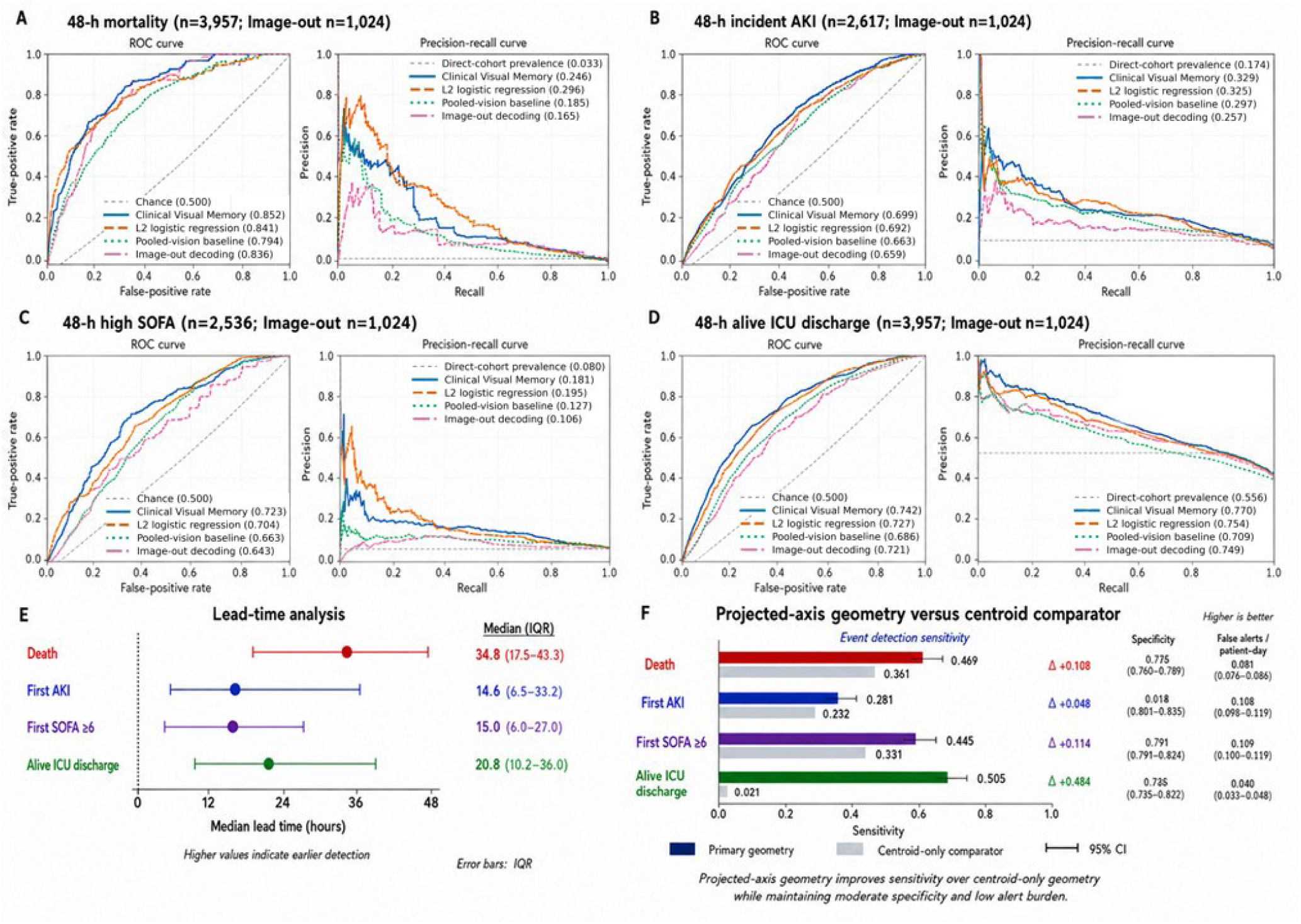
Held-out clinical prediction and critical-transition performance. (A-D) Receiver-operating-characteristic and precision-recall curves are shown for 48-hour mortality, incident AKI, high SOFA, and alive ICU discharge. Clinical Visual Memory is compared with L2-regularized logistic regression, a simpler pooled-vision baseline, and deterministic image-out decoding. Direct-prediction cohort sizes were 3,957 for mortality, 2,617 for AKI, 2,536 for high SOFA, and 3,957 for discharge; image-out decoding used 1,024 held-out stays per task. (E) Median lead time and IQR are shown for death, first AKI, first SOFA ≥6, and alive ICU discharge. (F) Event-detection sensitivity of projected-axis geometry is compared with centroid-only geometry; specificity and false alerts per patient-day are shown for the primary geometry. Error bars denote 95% CIs for sensitivity and IQRs for lead time, as indicated. Abbreviations: AKI, acute kidney injury; AUPRC, area under the precision-recall curve; AUROC, area under the receiver-operating-characteristic curve; CI, confidence interval; ICU, intensive care unit; IQR, interquartile range; SOFA, Sequential Organ Failure Assessment.

Neither AUPRC nor performance on the other three outcomes contributed to the reported retention percentage. Because restored images entered the encoder at unchanged dimensions, this experiment tested tolerance to loss of source-image information; it did not reduce DINOv2 patch-token count, memory use, or computational cost. This experiment explicitly tested the framework’s tolerance to extreme information density reduction while retaining generalist forecasting utility.

### Continuous forecasting via observed-state critical-transition geometry

To evaluate continuous clinical navigation capabilities, a separate critical-transition analysis utilized an observed eight-dimensional physiological-state space (cardiovascular, respiratory, renal, metabolic, hepatic, hematologic, neurological, and infection/inflammatory).

Critical-transition analyses were performed in a separate eight-dimensional clinical-state space comprising cardiovascular, respiratory, renal, metabolic, hepatic, hematologic, neurological, and infection/inflammatory domains. Each domain was represented on a bounded 0-to-1 deterioration scale at 3-hour resolution, with higher values indicating greater abnormality. For retrospective construction of training-set reference trajectories, interior missing observations could be linearly interpolated between observed values, but no extrapolation beyond the observed temporal range was performed. For held-out transition detection, the clinical state at time *t* was constructed using only information available at or before *t*; observations occurring after *t*, the eventual event time, and post-t outcomes were not used to calculate the transition score.

Four event clocks were analyzed: death, first incident AKI, first SOFA ≥6, and alive ICU discharge. For each event *q*, an event-state centroid *e_q_* was estimated from the eight-domain state at the event window. The empirical low-severity state *s_min_* defined the origin of an event-specific projected axis *a_q_* = *e_q_* − *s_min_*. Event-centroid distance and projected-axis position were calculated in the clinical state space. The projected-axis coordinate quantified progress from the low-severity state toward the event-associated state and was the primary geometry for held-out transition detection; centroid-only geometry served as the comparator. Event-state centroids, the low-severity reference state, event-specific axis vectors, and all standardization parameters were estimated exclusively from the training data and were fixed before evaluation of the held-out cohort. No held-out event labels were used to construct the event geometry.

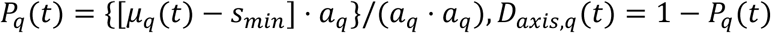

Approach velocity was defined as *V*(*t*) = −*dD*(*t*)/*dt* so that positive values represented movement toward the event state; acceleration was *A*(*t*) = *dV*(*t*)/*dt*. Distance, velocity, and acceleration were standardized, and the composite critical-transition index was defined as 0.50*z*[−*D*(*t*)] + 0.30*z*[*V*(*t*)] + 0.20*z*[*A*(*t*)]. For death, AKI, and high SOFA, increasing transition values represented deterioration; for alive ICU discharge, the same geometry was interpreted as movement toward recovery.

Operating thresholds for both the centroid-distance and projected-axis detectors were frozen independently on the training partition, before validation or held-out outcomes were examined. For each geometry, the threshold was set to the lowest or most sensitive score satisfying two prespecified training-set constraints simultaneously: stay-level specificity ≥ 0.80 and false alerts ≤ 0.10 per patient-day, evaluated over training-split non-event stays. Both detectors were thus governed by an identical dual-constraint operating criterion, a specificity floor combined with a false-alert-rate ceiling applied separately to each geometry’s own training score distribution. Thresholds were then applied unchanged to validation and test.

### Statistical analysis

Continuous cohort characteristics are reported as median and IQR, and categorical characteristics as counts and percentages. Preliminary descriptive comparisons used two-sided Mann-Whitney U tests for continuous variables and chi-square tests for categorical variables. Held-out discrimination was evaluated using AUROC and area under the precision-recall curve (AUPRC), with AUPRC included because outcome prevalence differed across tasks [35]. Calibration was summarized using the Brier score [36] and calibration slope. AUROC confidence intervals and paired between-model difference intervals were obtained through patient-level bootstrap resampling, preserving paired predictions from competing models. AUPRC confidence intervals were not available and numerical AUPRC differences were therefore not interpreted inferentially. Transition detectors were evaluated using sensitivity, specificity, false alerts per patient-day, and median warning lead time with IQR. Confidence intervals for transition metrics were calculated using 300 stay-level bootstrap replicates.

## RESULTS

### Cohort characteristics and multimodal data fragmentation

After exclusion of 19,629 ICU stays shorter than 24 hours, the eligible cohort included 54,551 patients, 68,546 hospital admissions, and 74,829 ICU stays (Figure 2). Baseline characteristics were summarized using the first eligible ICU stay per patient. Median age was 67 years (IQR, 55-77), 23,495 patients (43.1%) were female, and 5,881 (10.8%) died during the hospitalization. Patients who died were older than survivors (median, 72 vs 66 years) and had longer ICU stays (median, 93.4 vs 54.6 hours) (Table 1). Modality availability was highly fragmented, underscoring the necessity of a unified representational substrate. Structured EHR and vital-sign images were available for 54,543 (99.99%) and 54,548 (99.99%) index stays, respectively; clinical notes for 34,099 (62.51%); ECGs for 19,550 (35.84%); and chest radiographs for 6,661 (12.21%). All five modalities were simultaneously available for only 3,340 patients (6.12%), requiring explicit representation of modality availability rather than restriction to complete cases to achieve generalist forecasting.

**Table 1.**
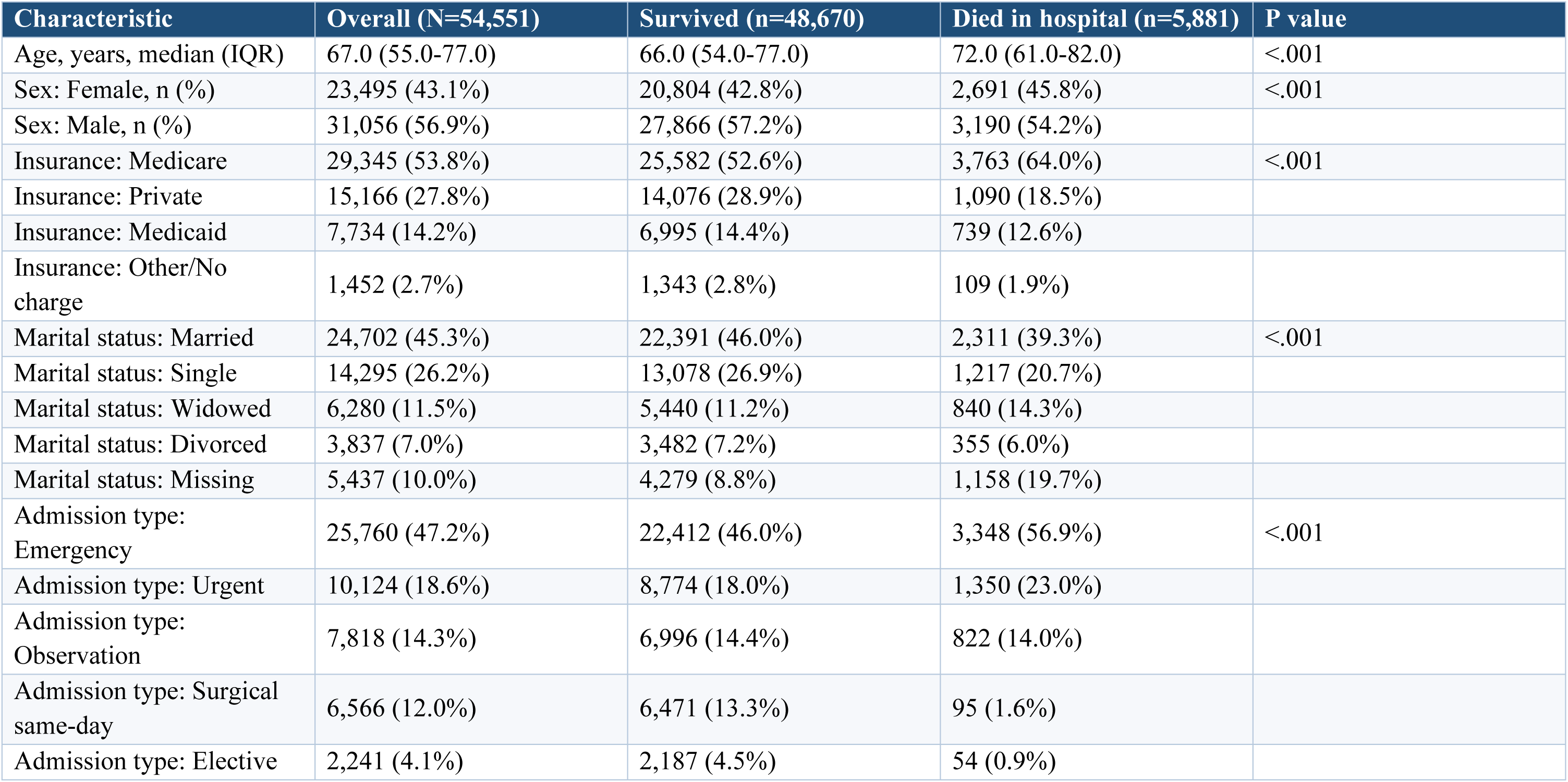
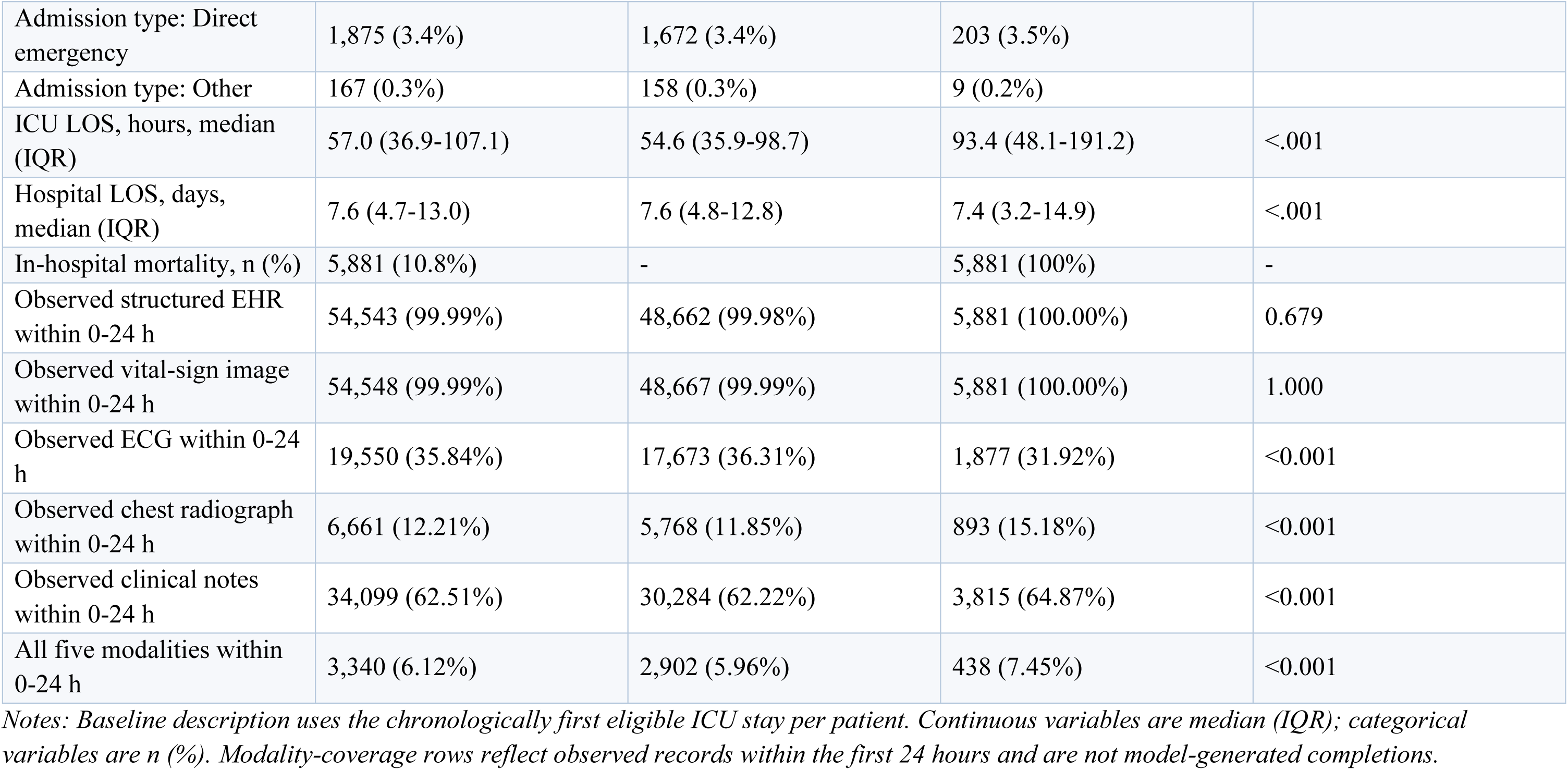
Baseline demographic and clinical characteristics of the index ICU stay per patient, MIMIC-IV adult cohort (ICU LOS ≥24 h), stratified by in-hospital mortality.

**Table 2.**
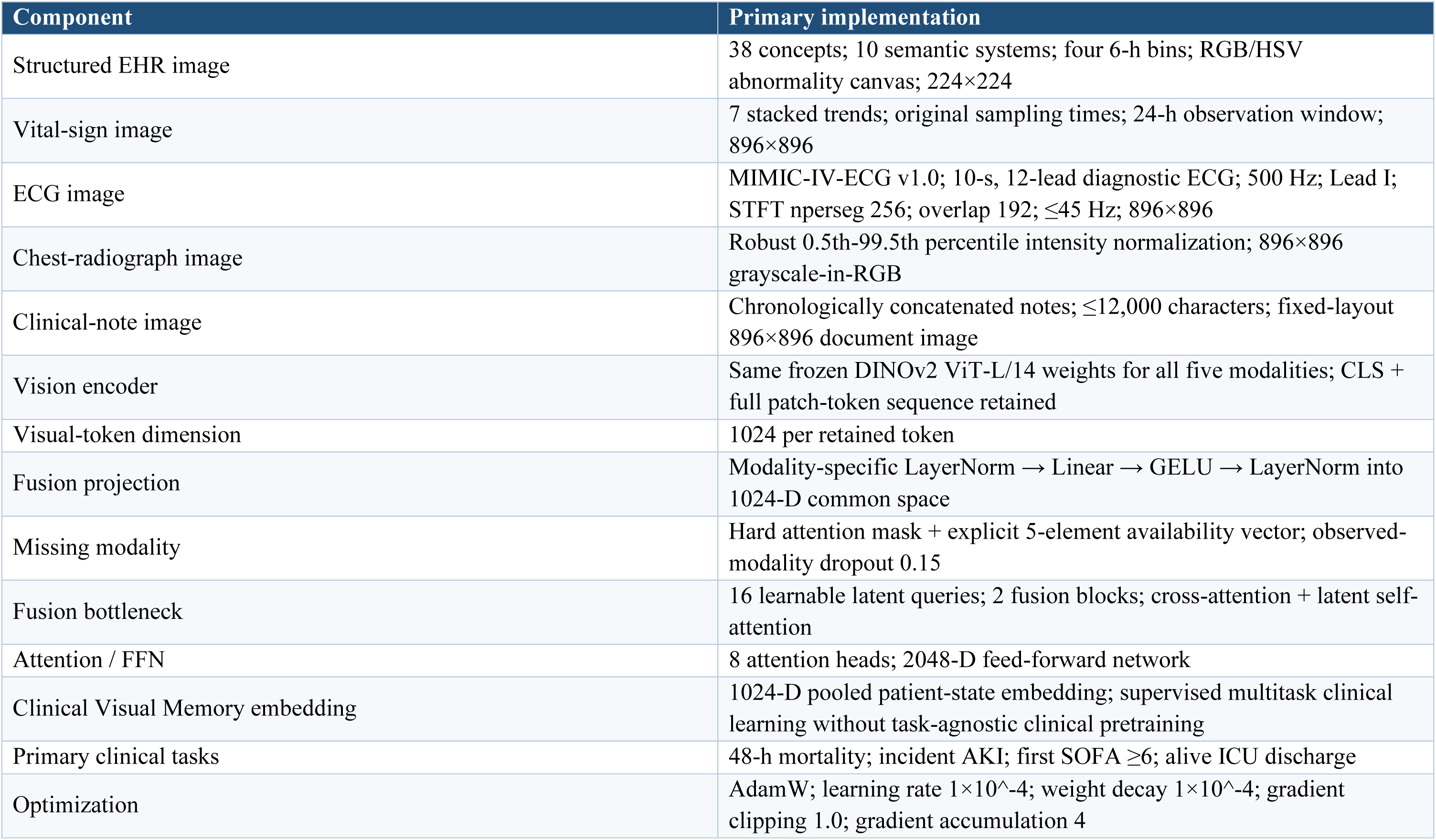
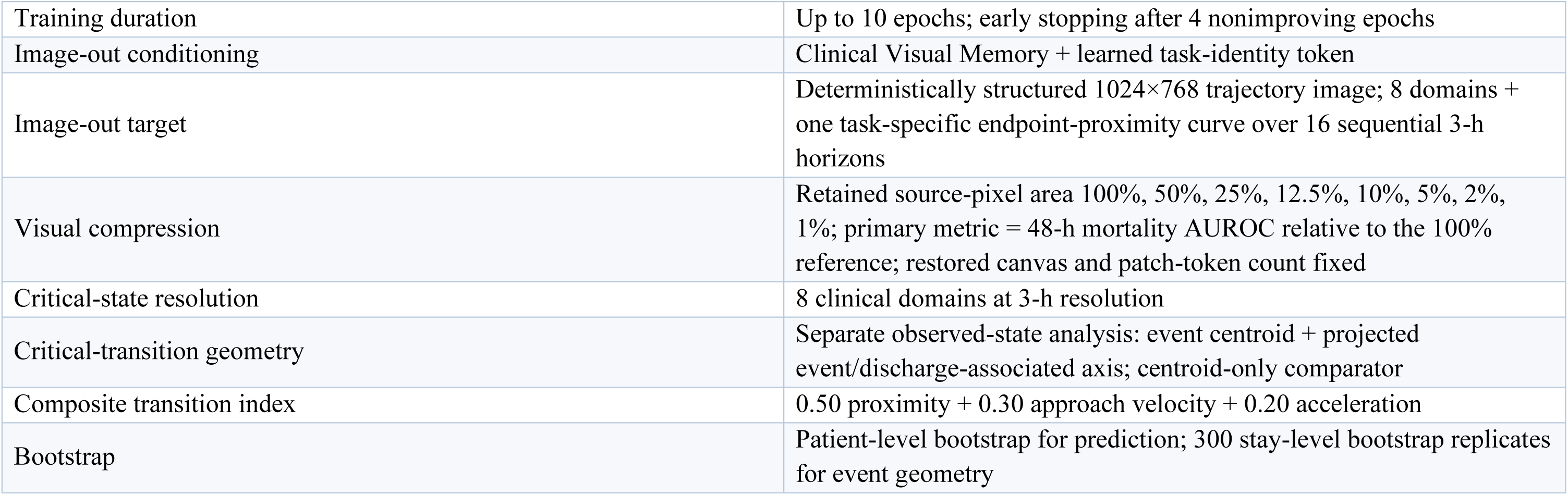
Primary architecture, training, compression, and critical-transition specification.

| Component | Primary implementation |
| --- | --- |
| Structured EHR image | 38 concepts; 10 semantic systems; four 6-h bins; RGB/HSV abnormality canvas; 224×224 |
| Vital-sign image | 7 stacked trends; original sampling times; 24-h observation window; 896×896 |
| ECG image | MIMIC-IV-ECG v1.0; 10-s, 12-lead diagnostic ECG; 500 Hz; Lead I; STFT nperseg 256; overlap 192; $\leq 45$ Hz; 896×896 |
| Chest-radiograph image | Robust 0.5th-99.5th percentile intensity normalization; 896×896 grayscale-in-RGB |
| Clinical-note image | Chronologically concatenated notes; $\leq 12,000$ characters; fixed-layout 896×896 document image |
| Vision encoder | Same frozen DINOv2 ViT-L/14 weights for all five modalities; CLS + full patch-token sequence retained |
| Visual-token dimension | 1024 per retained token |
| Fusion projection | Modality-specific LayerNorm → Linear → GELU → LayerNorm into 1024-D common space |
| Missing modality | Hard attention mask + explicit 5-element availability vector; observed-modality dropout 0.15 |
| Fusion bottleneck | 16 learnable latent queries; 2 fusion blocks; cross-attention + latent self-attention |
| Attention / FFN | 8 attention heads; 2048-D feed-forward network |
| Clinical Visual Memory embedding | 1024-D pooled patient-state embedding; supervised multitask clinical learning without task-agnostic clinical pretraining |
| Primary clinical tasks | 48-h mortality; incident AKI; first SOFA $\geq 6$; alive ICU discharge |
| Optimization | AdamW; learning rate $1 \times 10^{-4}$; weight decay $1 \times 10^{-4}$; gradient clipping 1.0; gradient accumulation 4 |
| Training duration | Up to 10 epochs; early stopping after 4 nonimproving epochs |
| Image-out conditioning | Clinical Visual Memory + learned task-identity token |
| Image-out target | Deterministically structured 1024×768 trajectory image; 8 domains + one task-specific endpoint-proximity curve over 16 sequential 3-h horizons |
| Visual compression | Retained source-pixel area 100%, 50%, 25%, 12.5%, 10%, 5%, 2%, 1%; primary metric = 48-h mortality AUROC relative to the 100% reference; restored canvas and patch-token count fixed |
| Critical-state resolution | 8 clinical domains at 3-h resolution |
| Critical-transition geometry | Separate observed-state analysis: event centroid + projected event/discharge-associated axis; centroid-only comparator |
| Composite transition index | 0.50 proximity + 0.30 approach velocity + 0.20 acceleration |
| Bootstrap | Patient-level bootstrap for prediction; 300 stay-level bootstrap replicates for event geometry |

### Validation of the unified visual-token memory

To establish the robustness of the compressed visual-token memory before testing its generative capabilities, we evaluated 48-hour prediction across four clinical tasks. The held-out task cohorts included 3,957 stays for 48-hour mortality, 2,617 for incident AKI, 2,536 for first SOFA score of 6 or greater, and 3,957 for alive ICU discharge. Operating as a generalist foundation, Clinical Visual Memory achieved AUROCs of 0.852 (95% CI, 0.821-0.882), 0.699 (0.673-0.725), 0.723 (0.688-0.757), and 0.742 (0.726-0.757), respectively (Figure 4A; Table 3). Corresponding AUPRCs were 0.246, 0.329, 0.181, and 0.770; Brier scores were 0.028, 0.133, 0.070, and 0.203; and calibration slopes were 1.008, 0.989, 0.970, and 0.972. The prespecified L2-regularized logistic-regression comparator achieved AUROCs of 0.841, 0.692, 0.704, and 0.727 for the four tasks. Paired 95% CIs for the AUROC difference, defined as comparator minus Clinical Visual Memory, were −0.051 to 0.030 for mortality, −0.033 to 0.019 for AKI, −0.063 to 0.025 for high SOFA, and −0.031 to 0.001 for alive ICU discharge; all included zero. Crucially, the pooled-vision comparator achieved significantly lower AUROCs of 0.794, 0.663, 0.663, and 0.686. Its paired differences from Clinical Visual Memory were −0.093 to −0.024, −0.057 to −0.015, −0.094 to −0.024, and −0.070 to −0.042, respectively, with all intervals strictly below zero, supporting the value of latent-query fusion relative to the evaluated pooled-vision representation. Precision-recall comparisons were task dependent. The logistic-regression comparator had numerically higher AUPRC for mortality (0.296 vs 0.246) and high SOFA (0.195 vs 0.181), whereas Clinical Visual Memory had numerically higher AUPRC for AKI (0.329 vs 0.325) and alive ICU discharge (0.770 vs 0.754). Confidence intervals were not available for AUPRC, precluding inferential comparisons.

**Table 3.**
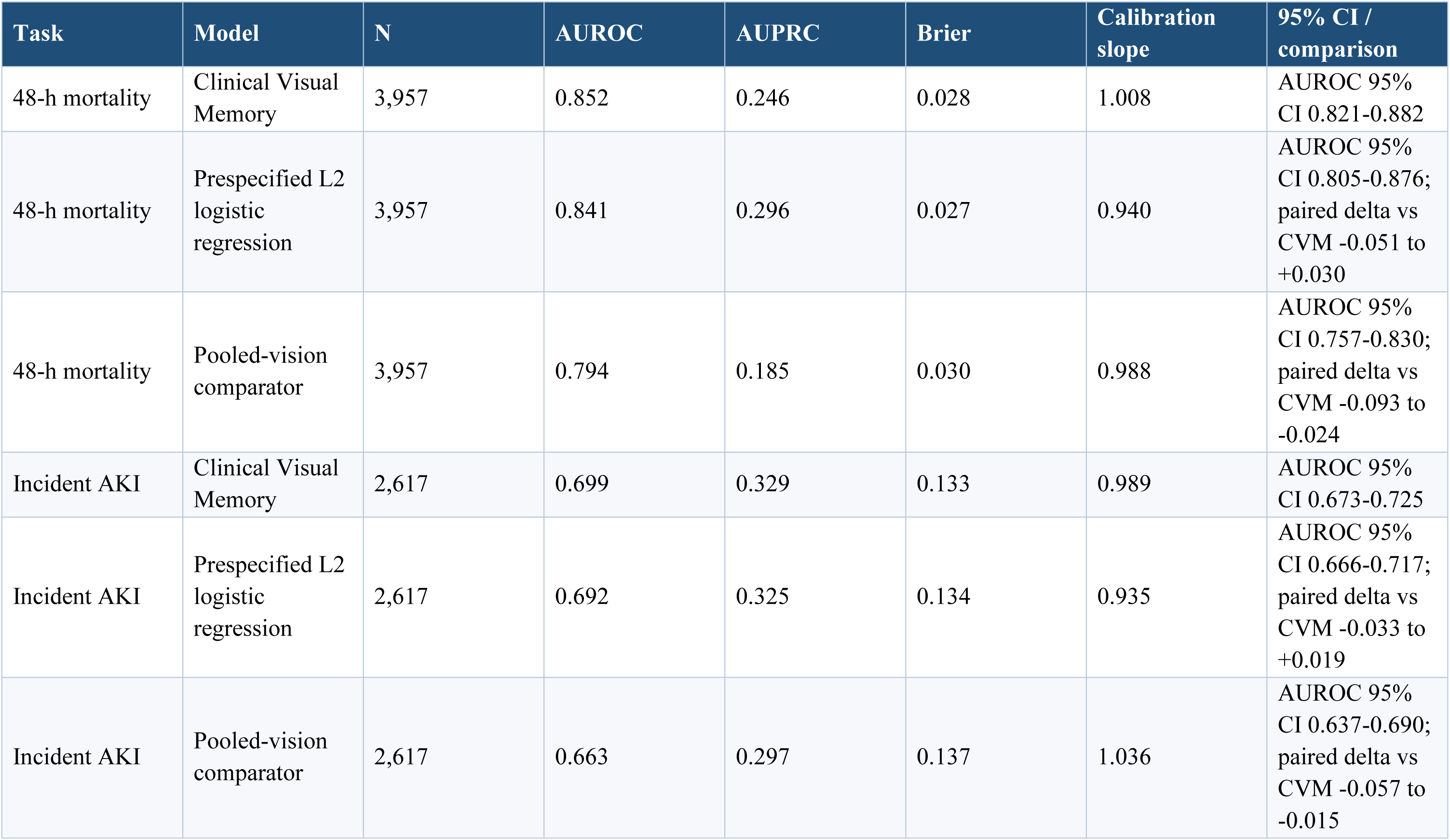
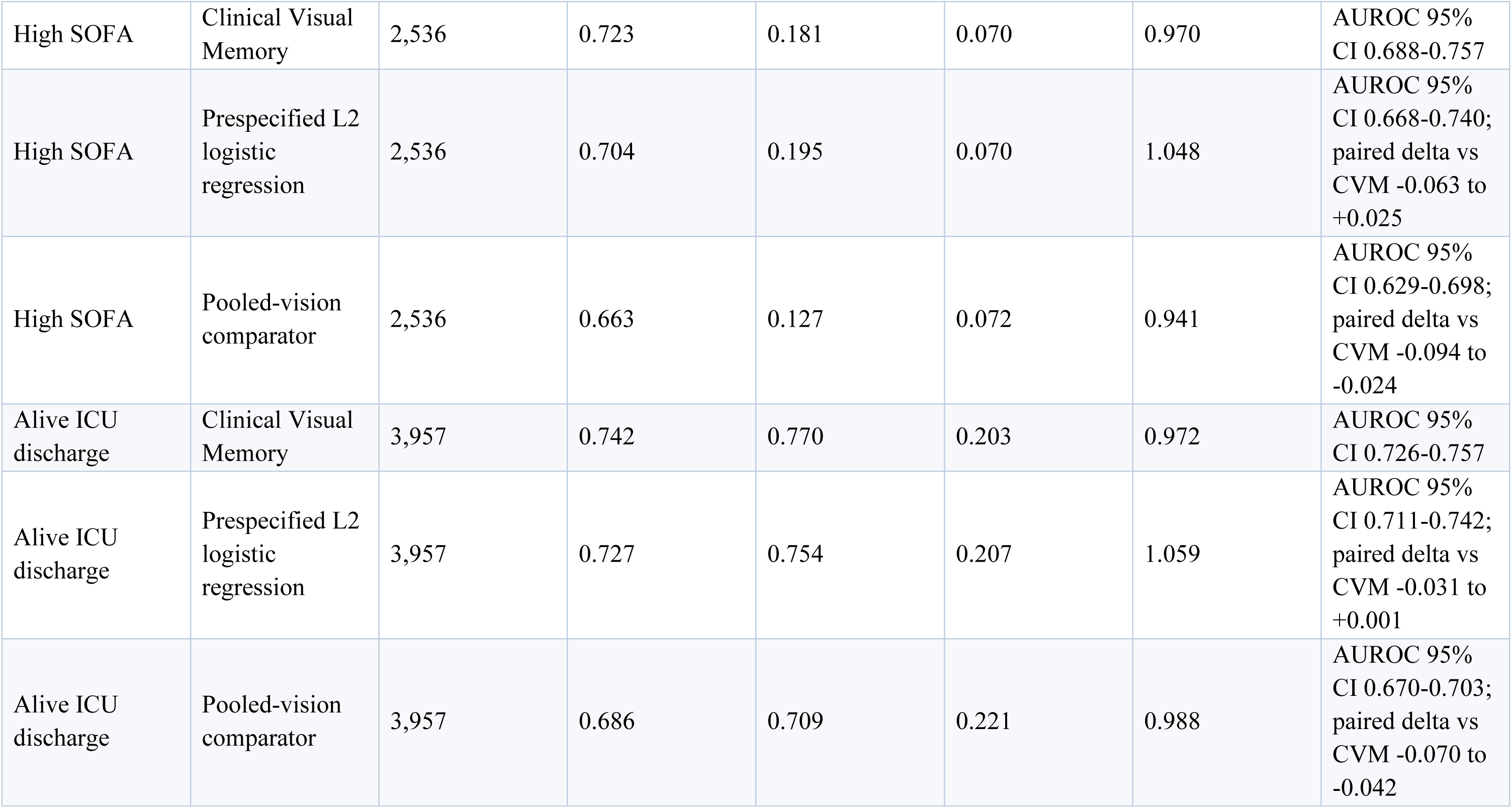
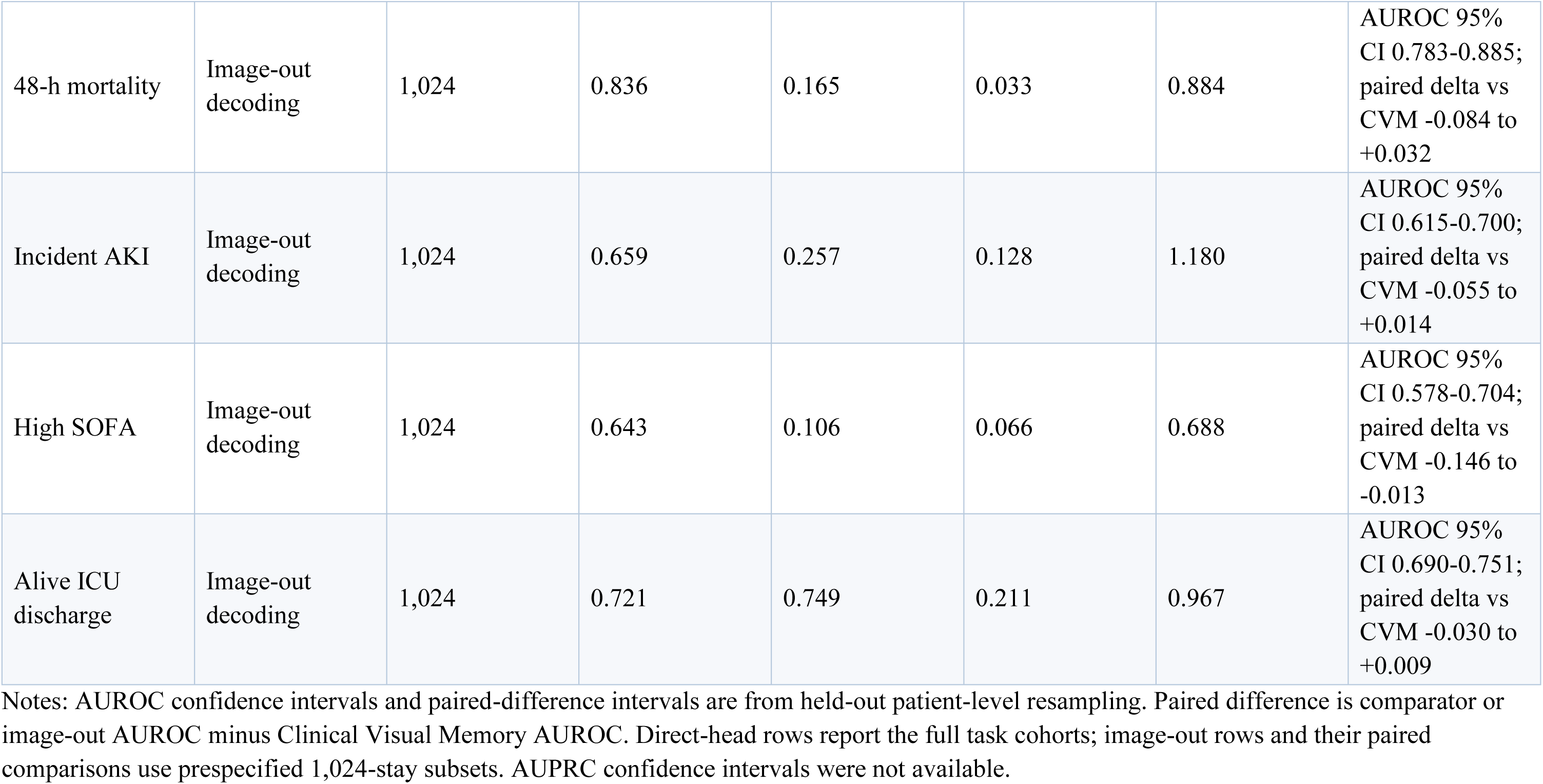
Held-out 48-hour multitask prediction, calibration, and image-out decoding.

| Task | Model | N | AUROC | AUPRC | Brier | Calibration slope | 95% CI / comparison |
| --- | --- | --- | --- | --- | --- | --- | --- |
| 48-h mortality | Clinical Visual Memory | 3,957 | 0.852 | 0.246 | 0.028 | 1.008 | AUROC 95% CI 0.821-0.882 |
| 48-h mortality | Prespecified L2 logistic regression | 3,957 | 0.841 | 0.296 | 0.027 | 0.940 | AUROC 95% CI 0.805-0.876; paired delta vs CVM -0.051 to +0.030 |
| 48-h mortality | Pooled-vision comparator | 3,957 | 0.794 | 0.185 | 0.030 | 0.988 | AUROC 95% CI 0.757-0.830; paired delta vs CVM -0.093 to -0.024 |
| Incident AKI | Clinical Visual Memory | 2,617 | 0.699 | 0.329 | 0.133 | 0.989 | AUROC 95% CI 0.673-0.725 |
| Incident AKI | Prespecified L2 logistic regression | 2,617 | 0.692 | 0.325 | 0.134 | 0.935 | AUROC 95% CI 0.666-0.717; paired delta vs CVM -0.033 to +0.019 |
| Incident AKI | Pooled-vision comparator | 2,617 | 0.663 | 0.297 | 0.137 | 1.036 | AUROC 95% CI 0.637-0.690; paired delta vs CVM -0.057 to -0.015 |
| High SOFA | Clinical Visual Memory | 2,536 | 0.723 | 0.181 | 0.070 | 0.970 | AUROC 95% CI 0.688-0.757 |
| High SOFA | Prespecified L2 logistic regression | 2,536 | 0.704 | 0.195 | 0.070 | 1.048 | AUROC 95% CI 0.668-0.740; paired delta vs CVM -0.063 to +0.025 |
| High SOFA | Pooled-vision comparator | 2,536 | 0.663 | 0.127 | 0.072 | 0.941 | AUROC 95% CI 0.629-0.698; paired delta vs CVM -0.094 to -0.024 |
| Alive ICU discharge | Clinical Visual Memory | 3,957 | 0.742 | 0.770 | 0.203 | 0.972 | AUROC 95% CI 0.726-0.757 |
| Alive ICU discharge | Prespecified L2 logistic regression | 3,957 | 0.727 | 0.754 | 0.207 | 1.059 | AUROC 95% CI 0.711-0.742; paired delta vs CVM -0.031 to +0.001 |
| Alive ICU discharge | Pooled-vision comparator | 3,957 | 0.686 | 0.709 | 0.221 | 0.988 | AUROC 95% CI 0.670-0.703; paired delta vs CVM -0.070 to -0.042 |
| 48-h mortality | Image-out decoding | 1,024 | 0.836 | 0.165 | 0.033 | 0.884 | AUROC 95% CI 0.783-0.885; paired delta vs CVM -0.084 to +0.032 |
| Incident AKI | Image-out decoding | 1,024 | 0.659 | 0.257 | 0.128 | 1.180 | AUROC 95% CI 0.615-0.700; paired delta vs CVM -0.055 to +0.014 |
| High SOFA | Image-out decoding | 1,024 | 0.643 | 0.106 | 0.066 | 0.688 | AUROC 95% CI 0.578-0.704; paired delta vs CVM -0.146 to -0.013 |
| Alive ICU discharge | Image-out decoding | 1,024 | 0.721 | 0.749 | 0.211 | 0.967 | AUROC 95% CI 0.690-0.751; paired delta vs CVM -0.030 to +0.009 |

Notes: AUROC confidence intervals and paired-difference intervals are from held-out patient-level resampling. Paired difference is comparator or image-out AUROC minus Clinical Visual Memory AUROC. Direct-head rows report the full task cohorts; image-out rows and their paired comparisons use prespecified 1,024-stay subsets. AUPRC confidence intervals were not available.

### Generative forecasting via task-conditioned image-out decoding

Completing the narrative arc from visual input to generative output, image-out decoding was evaluated in a prespecified random subset of 1,024 held-out stays per task. This generative forecasting interface produced AUROCs of 0.836 (95% CI, 0.783-0.885) for mortality, 0.659 (0.615-0.700) for AKI, 0.643 (0.578-0.704) for high SOFA, and 0.721 (0.690-0.751) for alive ICU discharge (Figure 4B; Table 3). Corresponding AUPRCs were 0.165, 0.257, 0.106, and 0.749; Brier scores were 0.033, 0.128, 0.066, and 0.211; and calibration slopes were 0.884, 1.180, 0.688, and 0.967. Paired 95% CIs for the AUROC difference between generative image-out decoding and the direct prediction head on the same selected stays were −0.084 to 0.032 for mortality, −0.055 to 0.014 for AKI, −0.146 to −0.013 for high SOFA, and −0.030 to 0.009 for alive ICU discharge. The intervals included zero for mortality, AKI, and discharge, demonstrating that the universal generative interface successfully translates the compressed visual memory into actionable clinical trajectories with no statistically clear difference detected for mortality, AKI, or discharge in the evaluated subsets. For high SOFA, the interval was entirely below zero, indicating lower discrimination through the generated-image pathway in the evaluated subset

### Optical compression fidelity under source-image information reduction

Validating optical compression as a highly efficient input mechanism capable of resolving multimodal data bottlenecks, mortality discrimination showed nonmonotonic tolerance to extreme source-image compression (Figure 3). Relative to the uncompressed 100%-area reference, retained AUROC was 104.3% at 50% retained source-pixel area, 86.3% at 25%, 98.9% at 12.5%, 62.4% at 10%, 46.5% at 5%, 21.2% at 2%, and 25.9% at 1%. Thus, at 12.5% retained area—a nominal 8-fold reduction in source-pixel area—the model retained 98.9% of its reference mortality AUROC. More aggressive reduction below this condition was associated with marked performance loss. Because images were restored to the original input canvas before encoding, encoder geometry, patch-token count, and computational sequence length remained unchanged. The experiment successfully confirmed the framework’s tolerance to massive loss of source-image information, proving that dense clinical data can be aggressively compressed while retaining generalist forecasting utility.

### Continuous forecasting via observed-state critical-transition geometry

To demonstrate continuous clinical navigation, a separate analysis of eight-domain observed clinical-state trajectories was conducted. Event-detection thresholds were selected on the training split under prespecified specificity and false-alert constraints and then applied unchanged to held-out data. For death, projected-axis geometry yielded sensitivity of 0.469 (95% CI, 0.427-0.516), specificity of 0.775 (0.760-0.789), and 0.081 false alerts per patient-day (0.076-0.086). Median lead time was 34.8 hours (IQR, 17.5-43.3) (Figure 4C-D; Table 4). Sensitivity was 0.108 higher than with centroid-only geometry (95% CI, 0.075-0.141). For first incident AKI, sensitivity was 0.281 (95% CI, 0.246-0.317), specificity was 0.818 (0.801-0.835), and false-alert burden was 0.108 per patient-day (0.098-0.119). Median lead time was 14.6 hours (IQR, 6.5-33.2), and the sensitivity difference from centroid-only geometry was 0.048 (95% CI, 0.007-0.088). For first SOFA score of 6 or greater, sensitivity was 0.445 (0.387-0.504), specificity was 0.808 (0.791-0.824), and false-alert burden was 0.109 per patient-day (0.100-0.119). Median lead time was 15.0 hours (IQR, 6.0-27.0), and the sensitivity difference was 0.114 (95% CI, 0.043-0.185). For the discharge-associated transition, projected-axis geometry yielded sensitivity of 0.505 (95% CI, 0.489-0.522), specificity of 0.778 (0.735-0.822), and 0.040 false alerts per patient-day (0.033-0.048). Median lead time before alive ICU discharge was 20.8 hours (IQR, 10.2-36.0). Sensitivity was 0.484 higher than with discharge-centroid-only geometry (95% CI, 0.467-0.501). These transition results validate that the underlying clinical-state space functions as a highly proactive navigation system rather than a static alert generator.

**Table 4.**
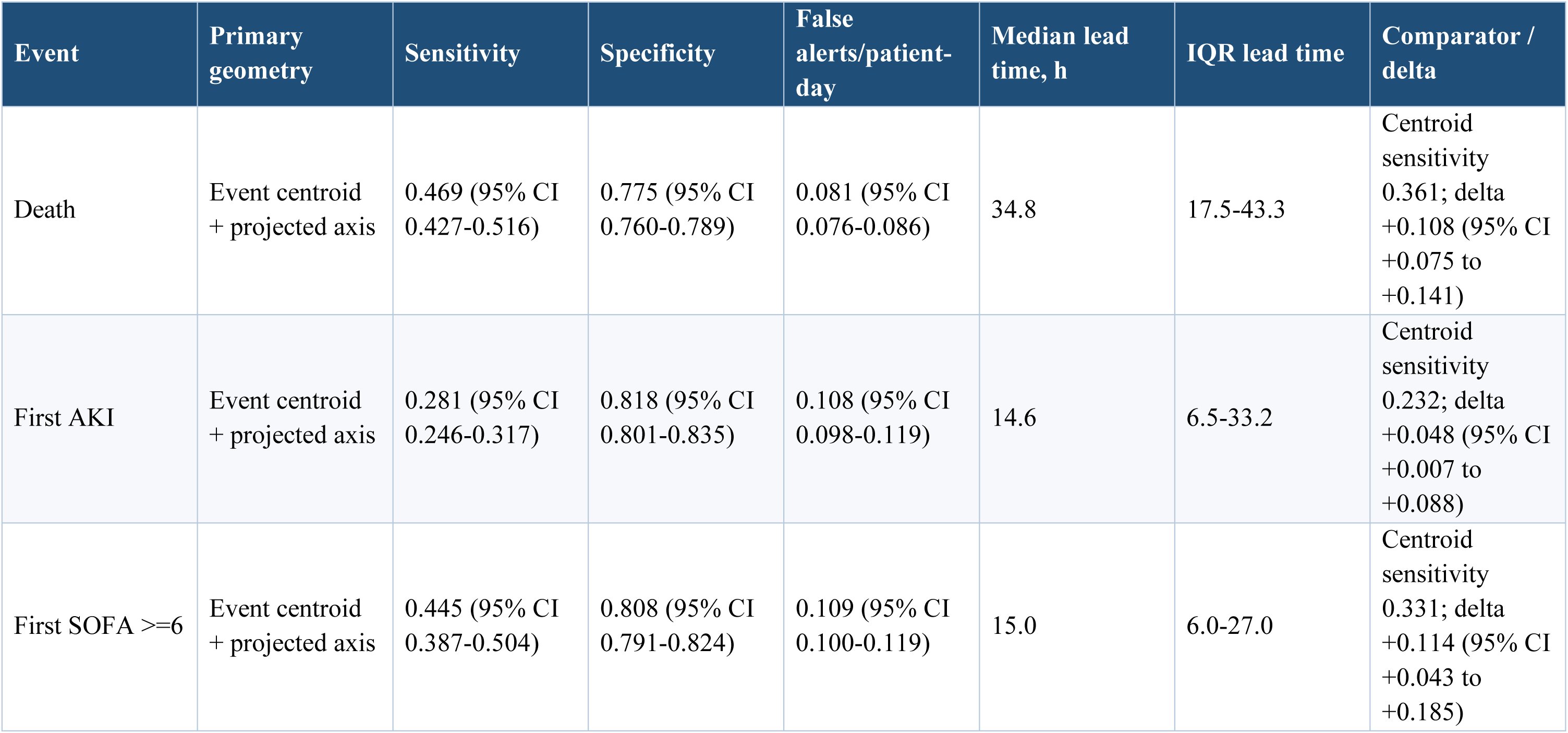
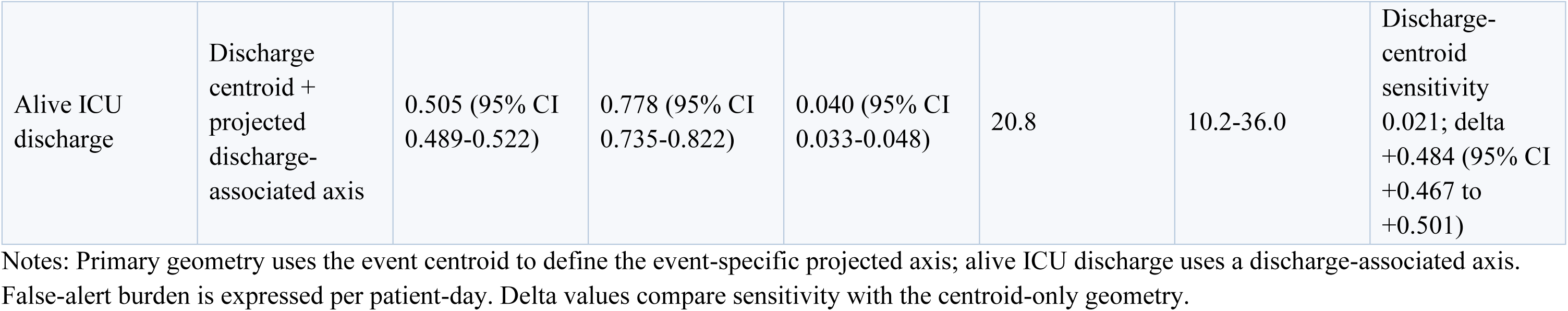
Held-out critical-transition detection and lead-time evaluation.

| Event | Primary geometry | Sensitivity | Specificity | False alerts/patient-day | Median lead time, h | IQR lead time | Comparator / delta |
| --- | --- | --- | --- | --- | --- | --- | --- |
| Death | Event centroid + projected axis | 0.469 (95% CI 0.427-0.516) | 0.775 (95% CI 0.760-0.789) | 0.081 (95% CI 0.076-0.086) | 34.8 | 17.5-43.3 | Centroid sensitivity 0.361; delta +0.108 (95% CI +0.075 to +0.141) |
| First AKI | Event centroid + projected axis | 0.281 (95% CI 0.246-0.317) | 0.818 (95% CI 0.801-0.835) | 0.108 (95% CI 0.098-0.119) | 14.6 | 6.5-33.2 | Centroid sensitivity 0.232; delta +0.048 (95% CI +0.007 to +0.088) |
| First SOFA $\geq 6$ | Event centroid + projected axis | 0.445 (95% CI 0.387-0.504) | 0.808 (95% CI 0.791-0.824) | 0.109 (95% CI 0.100-0.119) | 15.0 | 6.0-27.0 | Centroid sensitivity 0.331; delta +0.114 (95% CI +0.043 to +0.185) |
| Alive ICU discharge | Discharge centroid + projected discharge-associated axis | 0.505 (95% CI 0.489-0.522) | 0.778 (95% CI 0.735-0.822) | 0.040 (95% CI 0.033-0.048) | 20.8 | 10.2-36.0 | Discharge-centroid sensitivity 0.021; delta +0.484 (95% CI +0.467 to +0.501) |

Notes: Primary geometry uses the event centroid to define the event-specific projected axis; alive ICU discharge uses a discharge-associated axis. False-alert burden is expressed per patient-day. Delta values compare sensitivity with the centroid-only geometry.

### Computational efficiency of the compressed visual-token memory

To quantify the computational efficiency of the compressed visual-token memory, we conducted a hardware-anchored benchmark on a single NVIDIA A800 GPU with 80 GB of memory. Beginning with cached patch tokens from five modalities, the Clinical Visual Memory framework generated calibrated probabilities for four prediction tasks with a median latency of 7.57 ms per patient at a batch size of 1, while peak allocated GPU memory was 0.111 GiB. At a batch size of 32, amortized latency decreased to 0.645 ms per patient, corresponding to a throughput of 1,551 patients per second, with peak allocated GPU memory of 0.637 GiB. Although the simpler pooled-vision comparator achieved lower single-patient latency (1.95 ms), its throughput at a batch size of 32 was less than half that of the Clinical Visual Memory framework (655 versus 1,551 patients per second). The 15-epoch multitask fine-tuning stage required 73.0 minutes, whereas a 30-epoch replay of the downstream prediction head required 18.44 seconds. These measurements characterize the core cached-token-to-prediction pathway and exclude upstream DINO image encoding and downstream generative Image-Out decoding; they therefore should not be interpreted as end-to-end system latency or memory requirements.

## DISCUSSION

In this retrospective MIMIC-IV study, Clinical Visual Memory was evaluated as a comprehensive, paradigm-shifting framework that unites high-fidelity optical compression with generative clinical forecasting. Five highly heterogeneous modalities were optically compressed into a unified visual-token space, fused into one patient-state memory, and reused for direct prediction, task-conditioned trajectory generation, and controlled source-image compression.

Four findings define this contribution. First, latent-query fusion successfully resolved multimodal data bottlenecks, producing higher held-out AUROC than pooled visual features. Second, paired AUROC differences from the prespecified L2-regularized logistic-regression comparator included zero for every task, proving the visual memory matches mature tabular models while enabling generative functions they cannot provide. Third, at a tested eightfold reduction in source-pixel area, the full fusion model retained 98.9% of its uncompressed mortality AUROC, validating optical compression as an exceptionally efficient input mechanism. Fourth, task-conditioned image-out decoded this memory into actionable clinical trajectories, complemented by a separate observed-state trajectory geometry that supplied directional transition estimates and measurable lead time. Together, these results shift clinical AI from fragmented, task-specific predictive models to a generalist visual operating system.

The architecture resolves an important source of fragmentation and token bottlenecks in multimodal clinical artificial intelligence. Structured measurements, physiological trends, electrocardiography, radiography, and narrative documentation ordinarily require disparate preprocessing pipelines and modeling assumptions. Clinical Visual Memory applies the same frozen visual backbone to every rendered modality, while modality-specific projections preserve source identity and explicit masks distinguish unavailable data from observed normality. Sixteen latent queries then optically compress the available patch-token sequences into a unified 1024-dimensional memory. The consistent advantage over pooled vision suggests that cross-modal integration requires access to fine-grained visual tokens, which are seamlessly handled by this compression mechanism. While enabling a paradigm shift, these findings do not yet establish Clinical Visual Memory as a zero-shot clinical foundation model. The DINOv2 backbone was pretrained on nonclinical images, and the clinical projections, fusion layers, and prediction heads were trained through four prespecified supervised objectives without task-agnostic clinical pretraining. Furthermore, the comparisons do not establish superiority to tabular modeling: paired AUROC intervals merely did not identify a clear difference in the available samples, and logistic regression had higher AUPRC for mortality and high SOFA.

The more defensible interpretation is that optical compression into a visual-token memory achieves a numerical range of discrimination comparable to prespecified tabular comparators while unlocking a generalist generative forecasting interface those models cannot support. Demonstrating true foundation-model behavior will require transfer to unseen tasks, institutions, and modality combinations with limited task-specific retraining. Validating the input mechanism, source-image compression emerged as a core mechanistic result. At 12.5% retained source-pixel area, corresponding to eightfold nominal spatial compression, mortality discrimination was nearly preserved. Because images were restored to a common canvas, DINOv2 input geometry, tiling, and patch-token count remained fixed; the experiment instead proved that visual renderings of clinical data contain immense spatial redundancy that can be aggressively compressed without losing outcome-relevant information. This extends the optical-compression hypothesis from rendered text [18] to a highly complex, heterogeneous clinical setting. However, this compression curve was nonmonotonic, evaluated only through mortality AUROC, and lacked uncertainty intervals. Therefore, 12.5% retained area serves as an informative proof-of-concept operating point, motivating future modality-adaptive native token reduction.

Beyond preserving predictive fidelity, optical compression successfully resolves the severe computational bottlenecks that typically prohibit continuous, real-time multimodal AI in the ICU. By evaluating the core token-to-prediction pathway, we demonstrated that the compressed visual-token memory operates with extreme computational lightness: an amortized inference latency of 0.645 ms per patient and a peak allocated GPU memory of just 0.637 GiB at batch 32. Achieving a throughput of 1,551 patients per second on a single commercial GPU proves that this architecture is uniquely scalable. Rather than requiring massive, dedicated GPU clusters to process long clinical histories, this generalist visual operating system could feasibly monitor an entire health system’s ICU population in real-time. It is important to note that this specific benchmark captures the efficiency of the core memory-to-prediction pathway; full end-to-end latency—including the initial modality rendering, DINO encoding, and generative Image-Out diffusion steps—will require comprehensive future benchmarking. Nevertheless, these computational metrics confirm that transforming heterogeneous clinical data into a highly compressed visual-token memory fundamentally bypasses the token-crisis limitations of standard multimodal transformers.

The clinical output, generative forecasting, completes the framework’s narrative arc, testing whether the unified visual memory can support an expressive output grammar rather than only scalar risks. Each generated image acts as a continuous clinical navigation display, placing eight physiological domains beside an endpoint-proximity trajectory across sixteen sequential 3-hour horizons. In this sense, image generation provides a universal output interface, analogous to the broader generalist-vision principle described by Gabeur et al [19]. Using a “full-self-driving” metaphor, optical compression acts as the perception stack, while generative image-out functions as the navigation system. Within the 1,024-stay evaluation samples, paired AUROC differences between image-out decoding and direct prediction included zero for mortality, AKI, and alive ICU discharge, confirming the generative pathway successfully retains substantial endpoint information. High SOFA, however, showed measurable loss (calibration slope 0.688), requiring a deterministic patient-specific anchor with residual diffusion. This demonstrates that while generative forecasting is powerful, it must be carefully calibrated to ensure clinical decodability.

To further support continuous generative forecasting, the critical-transition analysis provided a complementary account of trajectory direction and velocity. In an eight-domain observed clinical-state space, projected-axis geometry improved sensitivity relative to centroid-only proximity for all four events. Median detection preceded death by 34.8 hours, AKI by 14.6 hours, high SOFA by 15.0 hours, and alive ICU discharge by 20.8 hours. The discharge-associated result demonstrates that this generative navigation system can map movement toward recovery as effectively as deterioration. These lead times do not automatically equate to bedside benefit, as sensitivity ranged from 0.281 to 0.505, false-alert burdens remained (0.081 to 0.109 alerts per patient-day), and endpoints like AKI and SOFA depend heavily on operational definitions and measurement frequency.

Several design features strengthen the study, including strict data partitioning, explicit modality-availability masking (addressing the reality that only 6.12% of index stays contained all five modalities), and out-of-sample threshold estimation. Important limitations include the retrospective, single-center nature of the MIMIC-IV data, which leaves transportability unknown. Furthermore, the four supervised endpoints do not prove zero-shot generalization, the tabular comparison was limited to one specification, and compression was evaluated without reducing actual patch-token computational costs. Most importantly, retrospective generative forecasting and warning lead times do not establish clinical actionability, safety, or improved patient outcomes. The framework was developed and evaluated at a fixed 24-hour landmark and therefore applies to patients remaining in the ICU at that time. Conditioning on this landmark excludes both early deaths and rapid ICU discharges, producing a case mix and event prevalence that differ from those of all ICU admissions. Generalist denotes a shared multimodal representation supporting multiple forecasting tasks; it does not imply validated invariance to observation-window duration. Performance before 24 hours or in stays ending earlier was not evaluated and should not be inferred from the present results.

The next stage must evaluate this paradigm-shifting framework as a generalist operating layer rather than a fixed risk model. This requires locked multicenter external validation, such as in eICU [37], transfer to unseen endpoints, native token reduction, and prospective silent deployment. If human-factors studies confirm clinicians can safely and accurately interpret this trajectory grammar, the fusion of optical compression and generative forecasting could replace dozens of fragmented data pipelines with a single, continuous visual-token navigation system.

## CONCLUSIONS

Clinical Visual Memory establishes a comprehensive, paradigm-shifting framework that unites high-fidelity optical compression with generalist generative forecasting. By mapping five heterogeneous ICU modalities into a unified visual-token memory, this architecture resolves critical multimodal data bottlenecks, notably retaining 98.9% of uncompressed mortality discrimination under a tested eightfold reduction in source-pixel area. Moving beyond the isolated scalar risks of standard predictive models—and achieving performance comparable to prespecified tabular comparators—this highly compressed, reusable representation successfully powers a continuous generative forecasting interface. Together with a separate observed-state geometry that supplies directional transition estimates and measurable warning lead times, these capabilities position Clinical Visual Memory as a transformative perception-and-navigation layer for critical care. While external validation, native token-efficiency testing, and prospective clinician-centered evaluation remain necessary before deployment, this end-to-end pipeline provides the definitive blueprint for a universal visual operating system in multimodal clinical intelligence.

## ADMINISTRATIVE INFORMATION

### Author Contributions

XBL had full access to all study data and takes responsibility for the integrity of the data and the accuracy of the data analysis.

**Concept and design**: XBL.

**Acquisition, analysis, or interpretation of data:** ZG, ZH, XBL.

**Drafting of the manuscript:** BJ, ZH, XBL.

**Critical revision of the manuscript for important intellectual content:** ZG, BJ, ZH, NO, TI, XBL

**Statistical analysis:** ZG, ZH, BJ, NO, TI, XBL.

**Administrative, technical, or material support:** XBL.

**Supervision:** XBL.

### Funding/Support

This work was supported by a sponsored research collaboration between Stanford University and Nippon Life Insurance Company of America; principal investigator: XBL.

### Ethics Declarations

This study was reviewed and approved by the Stanford University Institutional Review Board as part of a broader research protocol involving analysis of multiple clinical data sources. The present study used only de-identified data from MIMIC-IV, a publicly available critical-care database. The MIMIC-IV database was established with institutional review board approval from Beth Israel Deaconess Medical Center and the Massachusetts Institute of Technology, with appropriate authorization for secondary use of de-identified data. All analyses reported in this study were conducted in accordance with the Stanford IRB-approved protocol and the PhysioNet Credentialed Health Data Use Agreement. No new participants were recruited and no new human-subject data were collected specifically for the present analysis. This was a retrospective secondary-data analysis and not a prospective interventional study; therefore, clinical-trial registration was not applicable.

### Data Sharing Statement

This study used MIMIC-IV (version 3.1), MIMIC-IV-ECG: Diagnostic Electrocardiogram Matched Subset (version 1.0), linked MIMIC-CXR radiographs, and MIMIC-IV-Note clinical narratives, available through PhysioNet [21–24]. Access to credentialed MIMIC resources requires completion of applicable training and execution of the PhysioNet Credentialed Health Data Use Agreement. Per that agreement, patient-level data cannot be redistributed by the authors. Derived analytic code, model-training code, and non-patient-level artifacts, including evaluation scripts, will be made available upon publication.

## Data Availability

This study used MIMIC-IV (version 3.1), MIMIC-IV-ECG: Diagnostic Electrocardiogram Matched Subset (version 1.0), linked MIMIC-CXR radiographs, and MIMIC-IV-Note clinical narratives, available through PhysioNet [21-24]. Access to credentialed MIMIC resources requires completion of applicable training and execution of the PhysioNet Credentialed Health Data Use Agreement. Per that agreement, patient-level data cannot be redistributed by the authors. Derived analytic code, model-training code, and non-patient-level artifacts, including evaluation scripts, will be made available upon publication.

## Acknowledgments

We thank Mike Spaid and Isaac Fine for stimulating discussions during the preparation and filing of the Stanford University patent application related to this work.

## SUPPLEMENTARY TABLES

**Supplementary Table 1.**
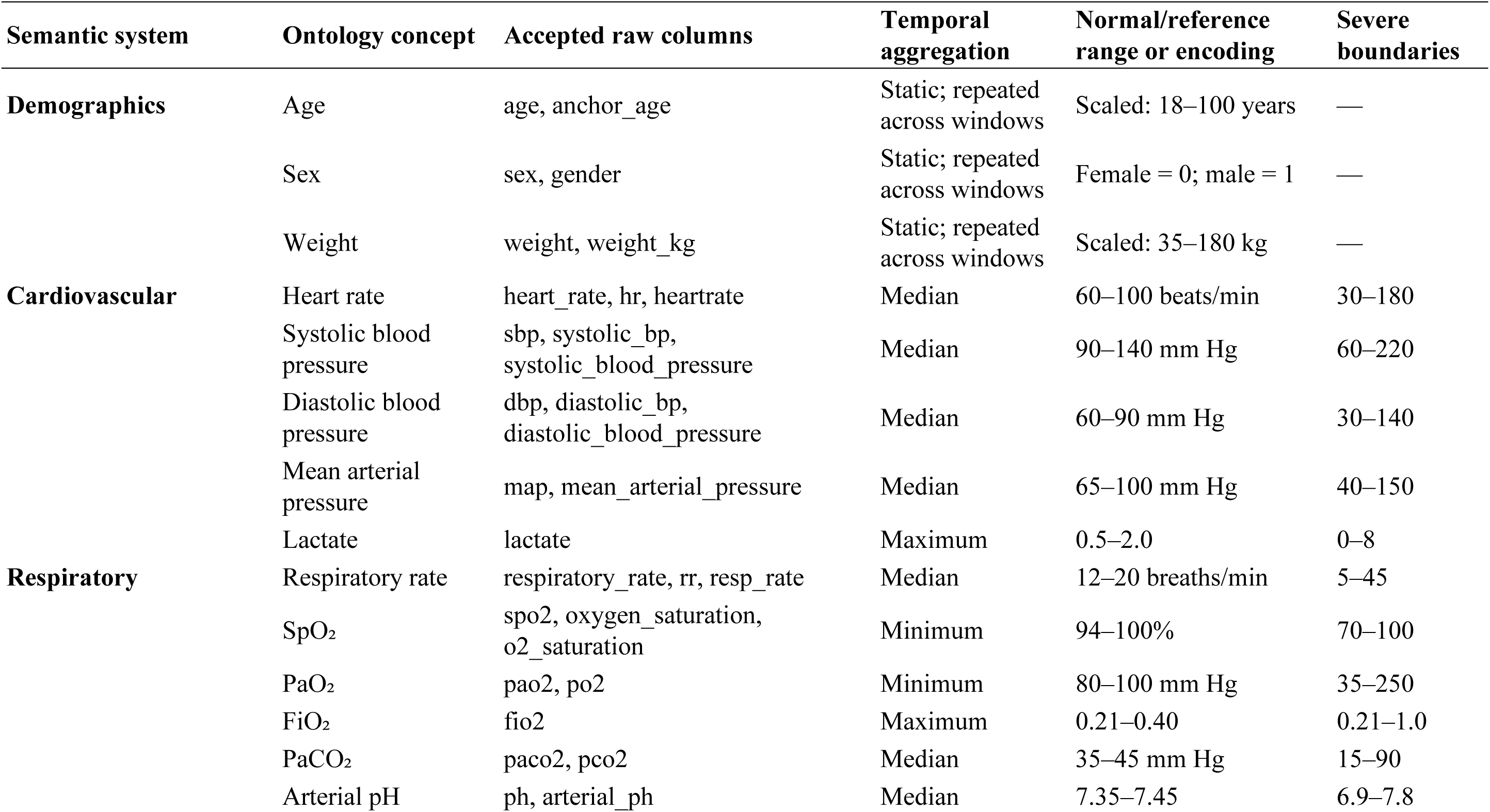
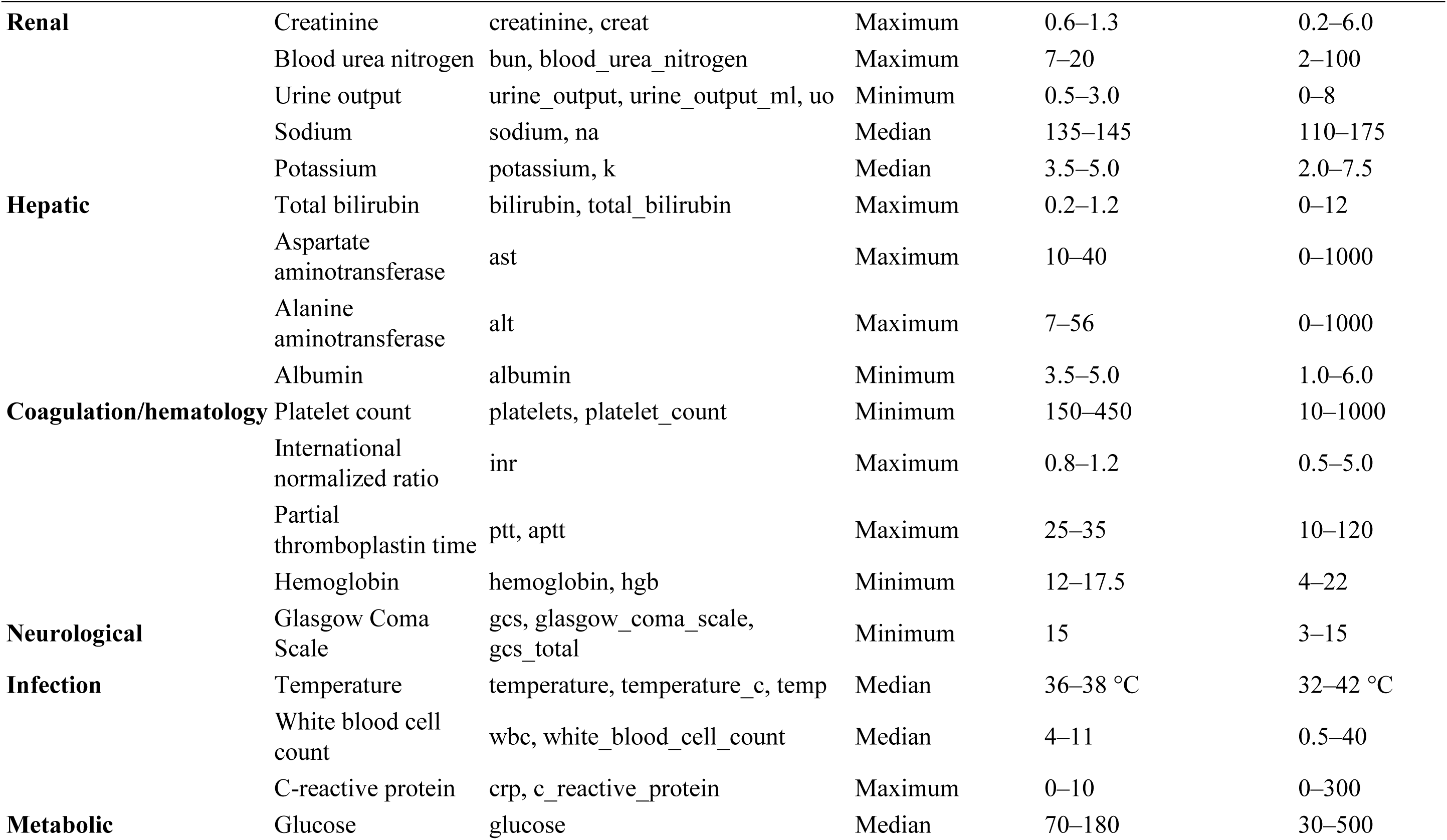
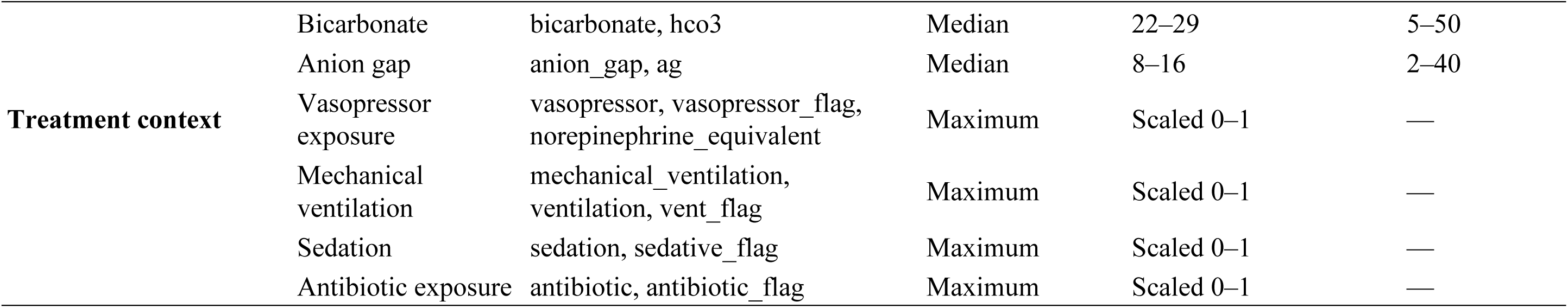
Structured EHR ontology concepts, input mappings, temporal aggregation, and encoding ranges.

